# Intensive Home-Based Motor Imagery Neurofeedback Training Using Mobile EEG to Assess Changes in Brain Activity and Motor Function in Stroke Survivors: A Multiple Baseline Design (MINTS Study Protocol)

**DOI:** 10.64898/2026.09.14.26363000

**Authors:** Jennifer Decker, Linda Bergmann, Stephanie Rosemann, Andreas Hein, Cornelia Kranczioch

## Abstract

**Background:** Motor imagery neurofeedback (MI-NF) engages sensorimotor brain networks and has shown potential for motor recovery after stroke. While laboratory-based studies have demonstrated neural and behavioral effects, evidence on real-world MI-NF training and its impact on brain activity and connectivity in stroke survivors remains limited. The Motor Imagery Neurofeedback Training in Stroke (MINTS) study investigates the effects of intensive, home-based MI-NF training using mobile EEG in chronic stroke survivors. Specifically, the study examines whether MI-NF training induces changes in neural activity and functional connectivity and whether these changes are accompanied by improvements in upper-limb motor function. Additionally, the study explores the temporal evolution of neural and behavioral changes across repeated baseline and training measurements.

**Methods:** MINTS is a prospective, single-group, multimodal intervention study using an across-subjects multiple-baseline design. Twenty-one individuals with chronic stroke (≥6 months post-stroke) and persistent upper-limb motor impairments undergo a pre-assessment phase, a variable-length baseline period, an intervention phase with 14 MI-NF training sessions, a post-assessment phase, and an exploratory three-month follow-up. MI-NF training is performed every second day at participants’ homes using mobile EEG. Repeated baseline and training measurements include grip strength, resting-state EEG, motor imagery tasks with and without neurofeedback, and daily-life movement monitoring. Pre- and post-assessments comprise comprehensive motor and cognitive testing, stationary EEG, and functional MRI. Primary outcomes are resting-state EEG coherence as an index of functional connectivity and MI-related EEG activity derived from mobile EEG within the multiple-baseline design. Secondary outcomes include MI-related EEG activity derived from stationary EEG, fMRI-based measures of brain activity and connectivity, clinical motor outcomes, and quantitative movement characteristics derived from movement sensors and video recordings. Tertiary outcomes and exploratory directions address broader motor and cognitive performance, associations between neural and behavioral changes, lesion-related effects, and persistence of training-related changes.

**Discussion:** By combining intensive home-based MI-NF training with repeated multimodal assessments, MINTS is designed to characterize neural and behavioral dynamics of motor recovery after stroke and to evaluate the potential clinical relevance of MI-NF training in everyday life settings.

**Trial Registration:** The study protocol was registered in the German Clinical Trials Register (DRKS00036147) on 25 March 2025.

## 1 Introduction

### 1.1 Background and Rationale

Stroke is a leading cause of long-term disability worldwide and frequently results in persistent motor impairments, particularly affecting upper-limb function, despite substantial advances in acute care and rehabilitation. While improvements in emergency treatment have increased survival rates, many stroke survivors require prolonged rehabilitation and nonetheless remain with chronic motor deficits that substantially limit independence and quality of life. This highlights the need for rehabilitation approaches that are not only effective, but also scalable, accessible, and suitable for long-term use in everyday life settings (1). Stroke-related motor impairments arise primarily from damage to central neural structures rather than peripheral musculature, suggesting that effective rehabilitation must target brain-level mechanisms of recovery. Neural plasticity describes the brain’s capacity to reorganize functional networks and establish new connections, including changes in large-scale functional connectivity patterns. It plays a central role in post-stroke recovery, however, the optimal strategy for harnessing this plasticity remains an open question. Contemporary models of post-stroke motor recovery increasingly conceptualize rehabilitation as targeting the reorganization of distributed motor networks across both hemispheres, including changes in ipsilesional and contralesional activity as well as their interhemispheric interactions (2–7).

Motor imagery (MI), defined as the mental simulation of a movement without overt execution, has emerged as a promising method for engaging motor-related neural networks after stroke (8). Neuroimaging and electrophysiological studies demonstrate substantial overlap between neural activity during MI and actual motor execution, particularly within sensorimotor and premotor regions (9, 10). By repeatedly activating these networks in the absence of physical movement, MI is thought to promote experience-dependent neural plasticity and support functional reorganization after stroke (11). MI can be performed without physical movement and therefore without the biomechanical load associated with repetitive motor training. Although MI may induce mental effort and cognitive fatigue in some individuals, it remains physically non-exhaustive and can be practiced frequently, making it especially suitable for individuals with limited motor capabilities.

The therapeutic potential of MI appears to be enhanced when combined with neurofeedback (NF), which provides real-time feedback on neural activity during training (12). NF enables participants to learn to modulate task-relevant brain activity in a targeted manner and may improve training engagement and specificity (13, 14). Electroencephalography (EEG)-based NF and brain-computer interface (BCI) approaches further offer a closed-loop framework that links motor intention to contingent sensory feedback, thereby promoting learning-dependent neural plasticity (15). In MI- and BCI-based stroke rehabilitation, feedback can be implemented through different modalities, including visual feedback, functional electrical stimulation, robotic assistance, or virtual reality-based feedback. Functional electrical stimulation appears particularly promising due to the additional proprioceptive and somatosensory feedback it provides (16), although meta-analytic evidence suggests that its effect size is not significantly higher than that of other BCI feedback modalities (17). These approaches represent complementary strategies with different technical requirements and potential therapeutic mechanisms. At the same time, visual feedback remains highly relevant, as it is commonly used in non-invasive EEG-based BCI systems (18, 19), has also been associated with improvements in upper-limb motor impairment after stroke (16) and offers a comparatively lightweight and scalable approach for repeated training outside controlled laboratory settings.

Clinical studies, systematic reviews, and meta-analyses indicate that MI-based NF and BCI-supported interventions can improve upper-limb motor outcomes after stroke and may induce neurophysiological changes related to motor recovery (12, 16, 18, 20–22). In parallel, neuroimaging and electrophysiological studies have shown that post-stroke motor recovery is associated with reorganization of distributed motor networks, including changes in ipsilesional and contralesional activity, interhemispheric interactions, and functional connectivity patterns (2–7). Functional connectivity measures have also been proposed as relevant biomarkers of cortical function and plasticity after stroke (23), and BCI studies have linked motor improvements to changes in ipsilesional activation, sensorimotor rhythm modulation, hemispheric lateralization, and structural integrity of motor pathways (18, 20, 24). Together, this literature supports the therapeutic and mechanistic relevance of MI-NF and BCI-based rehabilitation approaches, while also highlighting the need for studies that further characterize training response, long-term effects, optimal training protocols, and factors influencing individual outcomes (16, 18). Existing studies have often evaluated neural and behavioral effects primarily from pre-to post-intervention, which provides limited insight into how training-related neural changes evolve over the course of repeated training and how these changes relate to baseline variability, individual response patterns, and behavioral recovery. These questions are particularly relevant for home-based MI-NF, where intervention feasibility, training adherence, neural change, and motor outcomes need to be examined under conditions that closely resemble participants’ everyday lives. This is especially important for stroke survivors with motor impairments, for whom frequent travel to laboratory or clinical settings may be burdensome and may limit access to intensive rehabilitation. Previous work by Zich and colleagues (24) directly addressed this translational challenge by implementing intensive MI-NF training at participants’ homes using a wireless EEG system. In this small-scale study, three participants in the chronic phase after stroke completed a home-based MI-NF training protocol, which was associated with changes in motor performance as well as structural and functional brain measures. These findings provided initial evidence that intensive home-based MI-NF using mobile EEG is feasible and may support training-related neuroplastic changes, but the small sample size and case-report design limited broader conclusions regarding mechanisms, variability, and the relationship between neural and behavioral change.

The present study, Motor Imagery Neurofeedback Training in Stroke (MINTS), continues this line of research and builds on the same home-based^1^ MI-NF training protocol while extending it to a larger sample, a broader set of outcome measures, and an across-subjects multiple baseline design (MBD). The novelty of MINTS lies in the integration of three elements that have not yet been combined in this form in stroke rehabilitation research: intensive MI-NF training delivered in participants’ homes using mobile EEG, repeated MBD-based assessment of neural and motor changes across the training period, and multimodal characterization of brain activity, functional connectivity, clinical motor function, and everyday movement behavior. By bringing regular EEG-based MI-NF training into the participants’ everyday environment while simultaneously capturing neural and behavioral change from multiple complementary perspectives, MINTS aims to provide an informative framework for studying MI-NF-related recovery in chronic stroke survivors.

### 1.2 Objectives

The MINTS study has two primary objectives. The first primary objective is to examine whether MI-NF training is associated with systematic changes in resting-state functional connectivity, operationalized as resting-state EEG coherence, over the course of the training. The second primary objective is to examine whether MI-NF training is associated with systematic changes in MI-related brain activity independent of NF. This objective is assessed using two related EEG-derived measures during MI without NF: event-related desynchronization (ERD) or synchronization (ERS) and hemispheric lateralization of MI-related activity.

Secondary objectives are to further characterize the nature and localization of training-related neural changes. This includes examining alterations in task-related oscillatory brain activity, hemispheric lateralization patterns, and large-scale functional connectivity using pre- and post-training high-density EEG and functional MRI (fMRI) measures. By including modalities with high temporal and spatial resolution, the study seeks to determine where in the brain MI-NF-related changes occur and in what form, ranging from localized activation shifts to distributed network reorganization.

Beyond neural outcomes, the MINTS study aims to assess whether MI-NF training is associated with improvements in upper-limb motor function and everyday movements. To this end, standardized clinical motor assessments are combined with extended movement monitoring in daily life. The study design also allows us to evaluate whether neural changes, if observed, are accompanied by functionally meaningful improvements in everyday motor behavior.

An additional, tertiary objective is to describe participants’ subjective experience of intensive, home-based MI-NF training. By implementing the intervention in participants’ everyday environments, the study explores whether mobile EEG–based MI-NF training can be performed regularly over several weeks without imposing undue burden or disrupting daily routines. This information is intended to support the interpretation and future refinement of scalable and accessible neurorehabilitation approaches. Finally, the MINTS study is designed to generate exploratory insights into factors that may influence training response, including lesion characteristics and baseline neural measures.

Taken together, these objectives are guided by the overarching expectation that home-based MI-NF training will be associated with changes in resting-state functional connectivity, MI-related sensorimotor activity, and upper-limb motor performance. We further expect that training-related neural changes will be related to changes in behavioral motor outcomes. Because the MINTS study combines primary, secondary, tertiary and exploratory outcomes across several modalities, these expectations are specified at the protocol level, whereas more detailed analysis-level hypotheses will be formulated for the respective outcome analyses.

## 2 Methods and Analysis

### 2.1 Trial Design

The study employs an across-subjects MBD, a form of single-case experimental design in which repeated measurements are collected during a baseline phase of varying length before the intervention is introduced at staggered time points across participants (25). In this design, each participant serves as their own control (26), allowing within-subject changes over time to be examined while reducing the influence of external factors without requiring a separate control group. It is particularly suitable for small, heterogeneous clinical populations such as stroke survivors, where recruitment can be challenging and interindividual variability is high (25).

Baseline length ranges from three to eight assessment sessions and is randomly assigned within predefined lesion-location groups prior to study start (see Section 2.2.4 Randomization). This procedure ensures that no two participants within a lesion-location group share the same baseline length, resulting in a staggered onset of the intervention across individuals and thereby strengthening causal inference (27). In randomized MBDs, random assignment of baseline lengths replaces the requirement for a stable baseline as the primary mechanism for controlling time-related threats to internal validity (25, 27, 28). The staggered intervention onset is specifically intended to demonstrate that change occurs when and only when the intervention is introduced to a given participant, thereby distinguishing intervention-related changes from time-related effects or spontaneous recovery without requiring a stable pre-intervention state. Baseline variability and trends are therefore not criteria for determining intervention onset, but are instead explicitly incorporated into the statistical analysis by comparing within-phase slopes across baseline and intervention phases in addition to phase mean comparisons, rather than assuming a stable pre-intervention state. The unit of allocation is the individual participant. Following methodological recommendations (25, 29), each lesion-location group includes three participants. These participants form a triplet of three tiers, with each tier corresponding to one participant-level baseline/intervention series within the across-subjects MBD.

The MBD of this trial specifically applies to three repeatedly measured assessment components: resting-state EEG, MI without NF, and grip strength (see Section 2.4.3 MBD Baseline Measurements). Resting-state EEG is used to derive resting-state EEG coherence (see Section 2.5.1 Resting-State EEG Coherence), MI without NF is used to derive MI-related ERD/ERS and hemispheric lateralization (see Section 2.5.2 MI-Related EEG Activity (MBD, Mobile EEG)), and hand grip is used to derive grip strength (see Section 2.5.5 Motor Function and Movement Analysis). While the statistical power of small-sample designs is generally limited (30), several strategies are implemented to enhance internal validity, including multiple baseline assessments, consistent timing of measurements across all sessions, homogeneous inclusion and exclusion criteria, and grouping by lesion type (cortical, subcortical, or combined cortical/subcortical). Together, these measures ensure that the study can reliably assess potential effects of home-based MI-NF training despite inherent constraints in sample size.

The MBD measures are complemented by measures that are collected on at least two time points, but never continuously across baseline and intervention phase (see Section 2.3 Study Design). With this unique combination of MBD measures and more sparsely collected pre- and post-intervention measures, the study design supports meaningful analysis of training-related changes even if the target sample size cannot be fully achieved due to potential recruitment difficulties and dropouts.

### 2.2 Participant Population

A total of 21 stroke survivors will be included in this study. Included participants have chronic upper limb motor impairments and meet the eligibility criteria listed in Table 1. Eligibility is assessed in two steps.

**Table 1.** Inclusion and exclusion criteria.

| INCLUSION CRITERIA | EXCLUSION CRITERIA |
| --- | --- |
| ▪ Age: $\geq 18$ years<br>▪ First-ever, unilateral vascular event<br>▪ Chronic phase ($\geq 6$ months post-onset)<br>▪ Chronic motor impairment of upper limb following stroke<br>▪ Physical and mental suitability<br>▪ MRI suitability<br>▪ Resident in study region | ▪ Moderate to severe aphasia<br>▪ Apraxia<br>▪ Neglect<br>▪ Severe physical or psychological conditions beyond stroke<br>▪ No residual voluntary movement in the affected upper limb (self-reported motor function score = 0)<br>▪ Chronic sleep deprivation defined as a self-reported average sleep duration $< 4$ hours per night<br>▪ Drug use or central nervous system-impacting medication<br>▪ Concurrent participation in another stroke-related study |

First, interested individuals complete a telephone interview to collect preliminary information and screen for major ineligibility. During the telephone screening, they rate the motor function of each upper limb (including the arm, hand, and fingers) on a scale from 0 to 10, where 0 indicates no voluntary movement and 10 indicates no functional impairment. Individuals reporting a score of 10 for the non-affected upper limb and a score between 1 and 8 for the affected upper limb are considered eligible for further screening. Individuals reporting a score of 0 for the affected upper limb are excluded, as the study aims to investigate improvements in residual motor function and requires measurable upper limb motor performance. Upper limb impairment is characterized in greater detail during the study using standardized clinical motor assessments.

Second, individuals meeting the basic inclusion criteria are invited to the first in-person study session, where aphasia screening is conducted prior to any further assessments. Aphasia screening includes selected subtests of the Aachener Aphasia Test (31), namely the Token Test and the Written Language Test. Subtest performance is interpreted according to the Aachener Aphasia Test manual using the reported Stanine norms for aphasia severity. Participants are excluded if their performance corresponds to moderate or severe aphasia, defined as ≥22 age-corrected error points in the Token Test or a total score of ≤63 in the Written Language Test.

While MRI eligibility is considered during inclusion, unforeseen technical or participant-related factors, such as positioning constraints or discomfort, may still prevent successful MRI acquisition at a later stage. In such cases, participants remain included in the study and contribute to all non-MRI outcome measures. The inclusion and exclusion criteria were chosen to ensure that participants are likely to tolerate and benefit from intensive home-based MI-NF training, while enabling evaluation of intervention effects across a wide age range and diverse motor and cognitive profiles. Variability in impairment characteristics and time since stroke onset (≥ 6 months post-event) is expected to provide insights into inter-individual differences in long-term recovery and adaptation.

To take into consideration the aspect of differences in brain injury patterns already in the trial design, in particular the across-subjects MBD aspect, participants are assigned to a lesion-location group before the intervention begins. Grouping is based on MRI findings obtained during the second study session. A neuroradiologist reviews the T1- and FLAIR-weighted scans to determine lesion location and assigns participants to one of three groups: cortical, subcortical, or combined cortical/subcortical.

#### 2.2.1 Sample Size

The target sample size of 21 participants was determined based on feasibility and statistical considerations. Because comparable studies involving similar interventions, outcome measures, participant groups, and study designs are currently lacking, reliable a priori effect size estimates for the primary outcomes were not available. Consequently, a formal outcome-specific sample size calculation was not feasible. Instead, sensitivity analyses were conducted using G*Power (32) to determine the minimum effect sizes detectable with the planned sample size across the principal statistical approaches used in the study. With *α* = 0.05 and power = 0.80, a sample of *N* = 21 allows detection of medium-to-large effects (*dz* = 0.65) in paired *t*-tests (two-tailed) and medium effects (*f* = 0.27) in repeated-measures ANOVA (*F*-tests). For secondary analyses involving pairwise correlations, directly comparable studies were likewise unavailable. However, Wu, Quinlan (23) reported correlations between EEG-based connectivity measures and improvements in hand function following a 28-day physical motor therapy training. For a sample size of *N* = 12 participants in the chronic stage of stroke recovery, the reported significant effects corresponded to correlation coefficients ranging from *r* = 0.58 to *r* = 0.77. In line with this range, sensitivity analyses using the same parameters as above indicated that effects of approximately *r* = 0.57 (medium to large) can be detected with our planned sample size.

While the sample size may limit the detection of smaller effects, it provides adequate sensitivity for identifying clinically meaningful and neurophysiologically relevant changes and supports the study’s aims. In the case of a very small final sample or incomplete lesion-location groups, visual analyses can also be applied to individual participants’ datasets, and the inclusion of multiple baseline measurements in the experimental design allows for improved interpretation of intervention-related changes.

#### 2.2.2 Recruitment

Participants will be recruited continuously until the end of the study, with more active recruitment periods initiated as needed to maintain the target sample size. Recruitment follows a multi-pronged approach, including newspaper articles in regional media, through partner clinics (Klinikum Bremen-Ost and Rehazentrum Oldenburg), presentations at local stroke support groups, and public information events at the University of Oldenburg. Initial contact is established either by interested individuals responding to study advertisements or by members of the study team following recruitment events or presentations. During initial contact, the first telephone interview is conducted (see Section 2.2.3 Timeline), and information on potentially eligible participants is entered into Castellum (33). Castellum is a secure, General Data Protection Regulation (GDPR)-compliant, open-source web application for participant recruitment and management that supports the organization of contact information, study participation, appointments, and pseudonymized study workflows.

#### 2.2.3 Timeline

The study procedures follow a standardized sequence for all participants (see Figure 1). Basic demographic and medical information are collected in the first phone call, and a detailed eligibility screening is performed, covering stroke history and medical background, in a second phone call. Once study capacity allows, the participant database is reviewed, and individuals meeting inclusion criteria without indications of exclusion factors are contacted. During a third call, study procedures are explained in detail, and appointments are scheduled (see Table 2).

**Figure 1:**
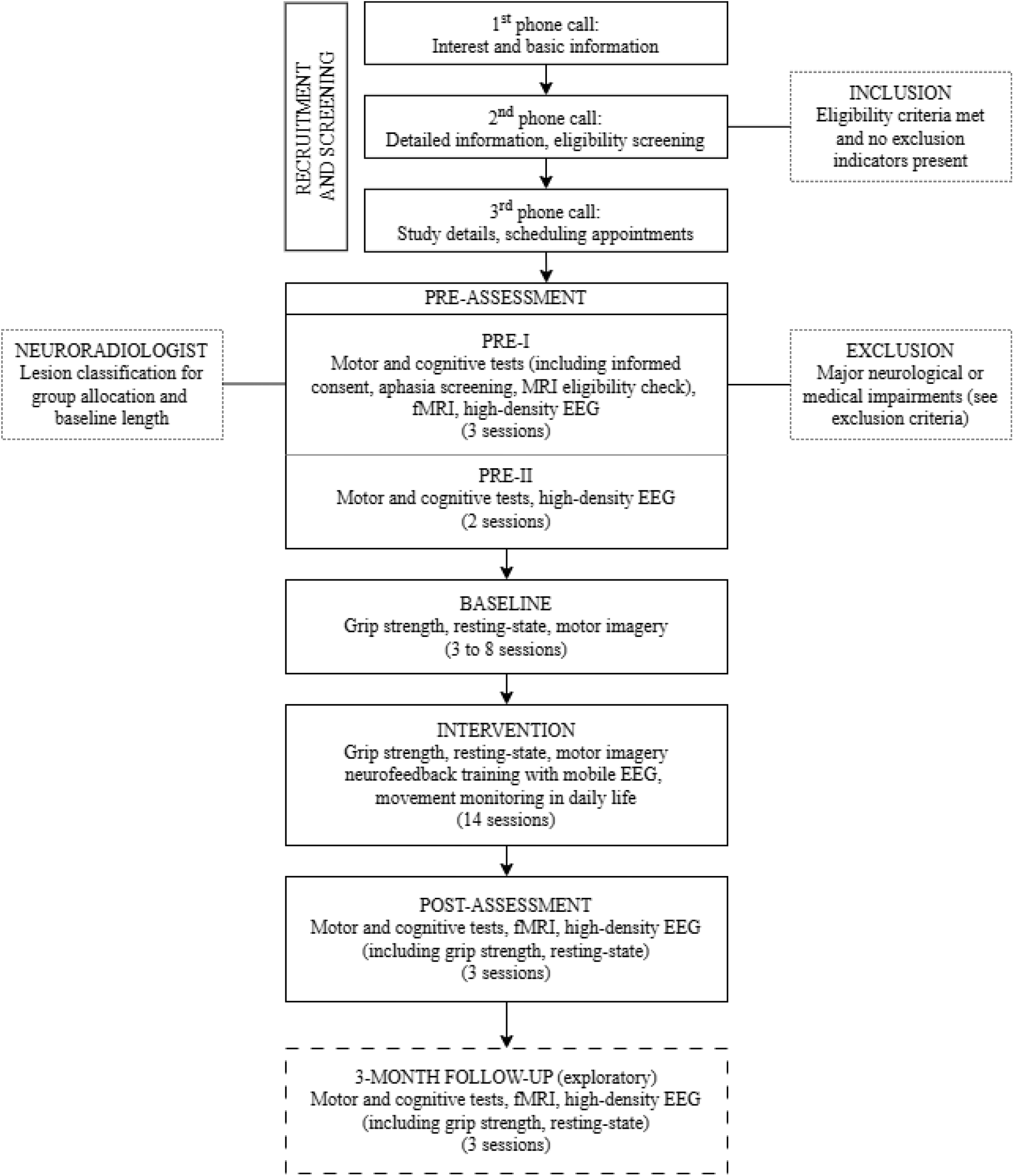
Study timeline. Schematic overview of recruitment and screening procedures, pre-assessment, baseline phase, intervention (MI-NF training), post-assessment, and exploratory three-month follow-up. Motor, cognitive, EEG, and MRI assessments are conducted at pre- and post-assessment, with grip strength, resting-state EEG, and motor imagery measured repeatedly during baseline and intervention.

**Table 2:**
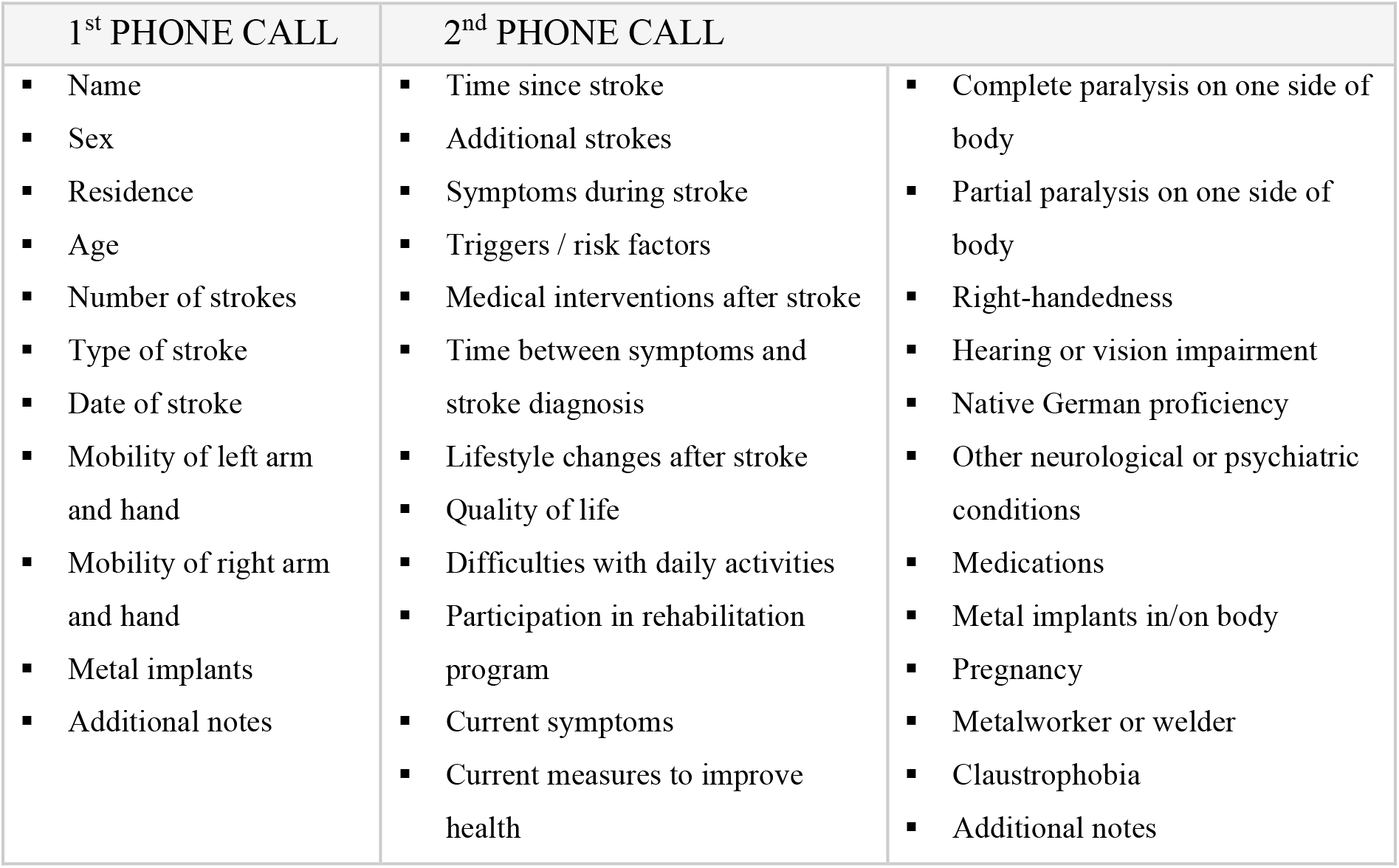
Participant information collected in the first and second telephone calls.

The pre-assessment sessions include standardized motor and cognitive tests, stationary high-density EEG recordings, and structural and functional MRI, including a motor execution/attempt task. During the motor tests, movements are additionally documented using synchronized video recordings and wearable motion sensors. If severe neurological impairments or other exclusion criteria are identified during screening or testing at the first study session, the individual is not enrolled further. MRI scans are acquired in a second session, and structural MRI data are reviewed by a neuroradiologist, who classifies stroke-related lesions and assigns participants to one of the three lesion-location groups. Based on this classification, participants are allocated within their respective lesion-location group according to the triplet-based randomization procedure described below (see Section 2.2.4 Randomization), which determines the number of baseline sessions completed before intervention onset.

The intervention phase comprises 14 training sessions over four weeks, consisting of MI supported by visual NF, conducted at the participants’ homes. In addition to the NF training, movements during selected activities of daily living are recorded using wearable sensors to capture changes in everyday motor behavior. Post-assessment sessions repeat all pre-assessment measures. Participants who consented to follow-up are invited for an additional assessment approximately three months after the post-assessment, during which all post-measurements are repeated.

#### 2.2.4 Randomization

To enable analyses of the MBD outcomes both across and within lesion-location groups, participants are allocated using a triplet-based randomization procedure (see Figure 2). Prior to participant enrollment and the start of data collection, a custom MATLAB script generated seven triplets, each consisting of three unique baseline lengths ranging from three to eight sessions. Within a triplet, no baseline length occurs more than once. Following MRI acquisition and neuroradiological review, participants are classified into one of three lesion-location groups (cortical, subcortical, or combined cortical/subcortical). Participants are then assigned to the next available position within an open triplet corresponding to their lesion-location group. The assigned position determines the participant’s baseline length and, consequently, the timing of intervention onset. If no open triplet exists for a participant’s lesion-location group, a new triplet is opened and assigned to that group, provided that fewer than seven triplets have been opened. Since the maximum sample size is 21 participants, no additional triplets will be opened once all seven triplets have been initiated. If, at this stage, a participant cannot be assigned to an open triplet matching their lesion-location group, the participant will be allocated to an incomplete triplet regardless of lesion-location group. This procedure ensures that participants can still be allocated to complete triplets, which is required for the randomization-based analyses of the MBD outcomes (see Section 2.6.1 Statistical Methods). As a consequence, up to two triplets may include participants from more than one lesion-location group. Such mixed triplets will not be used for exploratory analyses of lesion-location effects (see Section 2.5.8 Exploratory Directions), but remain valid for the MBD-based analyses because baseline-length randomization within the triplet is unaffected by lesion-location composition. As participant enrollment and lesion characteristics cannot be predicted in advance, the number of triplets allocated to each lesion-location group and the resulting group sizes cannot be specified a priori and may differ across groups. This allocation procedure is required for the MBD (34), in which participants begin the intervention after varying baseline durations. Staggering intervention onset across participants helps control for time-related effects and strengthens causal inference regarding intervention-related changes. Random assignment of baseline lengths within each lesion-location group reduces predictability of intervention timing and minimizes potential allocation bias while maintaining an overall average baseline duration of approximately 5.5 sessions across the study. The randomization sequence was generated before recruitment commenced, stored digitally, and referenced throughout participant allocation to ensure adherence to the predefined stratification procedure.

**Figure 2:**
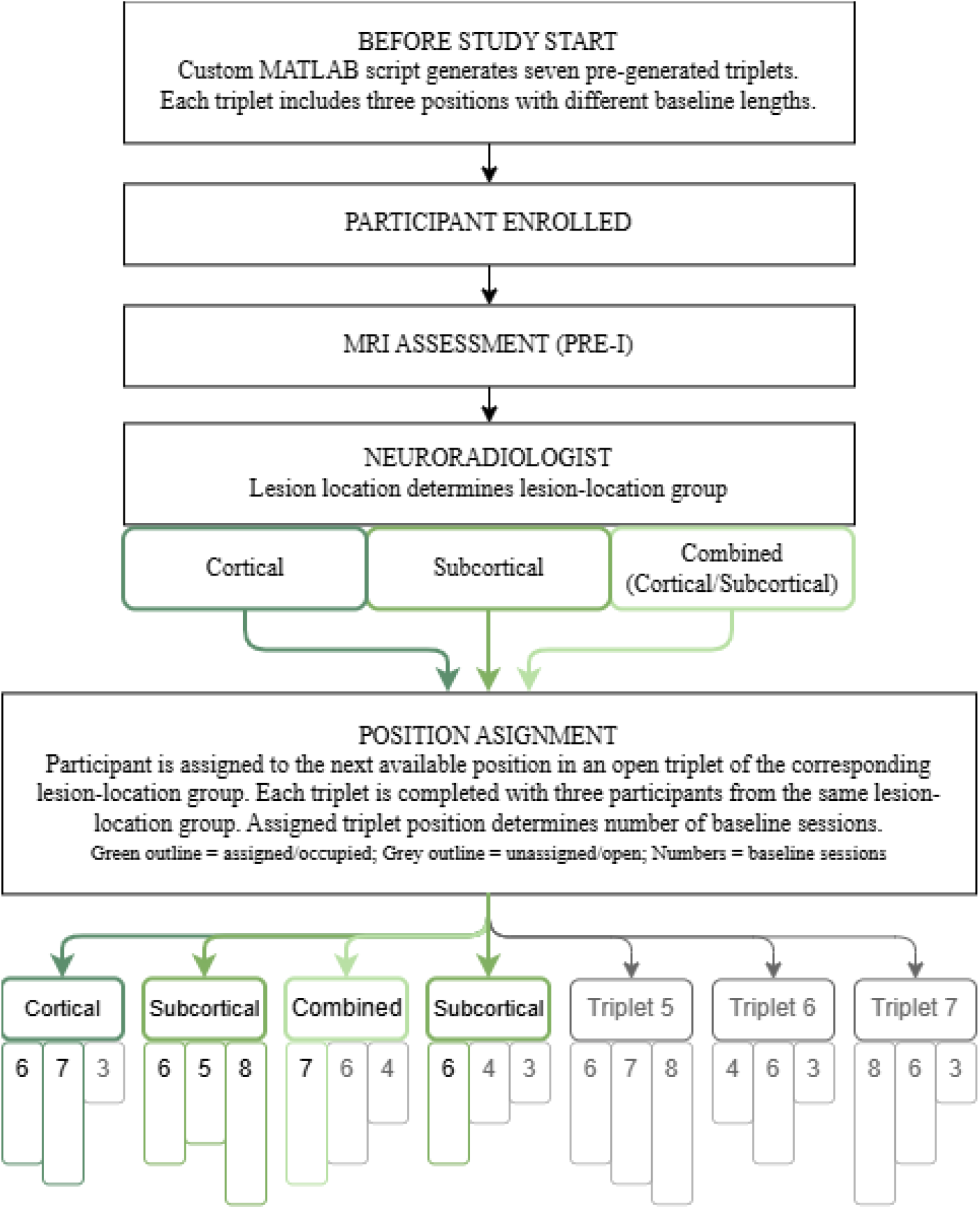
Triplet-based randomization procedure used for baseline-length allocation. Participants are first classified into cortical, subcortical, or combined cortical/subcortical lesion-location groups based on MRI findings. Within each lesion-location group, participants are sequentially assigned to the next available position in a pre-generated triplet. The first participant assigned to a triplet determines the lesion-location category of that triplet, and the remaining positions are subsequently filled by participants from the same lesion-location group whenever possible. Each triplet consists of three positions with distinct predefined baseline lengths selected from a range of three to eight sessions, and the assigned position determines the timing of intervention onset. The lesion-location assignments and baseline lengths shown are illustrative examples only and do not represent the actual randomization sequence used in the study.

### 2.3 Study Design

The MINTS study follows a structured, multi-phase design consisting of a pre-assessment phase, a variable-length baseline period, a four-week home-based intervention (MI-NF training), and a post-assessment phase with an exploratory three-month follow-up. Figure 3 illustrates the sequencing and timing of all session types using the example of a participant who completes six baseline assessments. The overall structure and order of sessions remain identical across participants; only the total study duration varies, ranging from a minimum of 23 sessions over approximately 6.5 weeks (three baseline assessments) to a maximum of 28 sessions over 8 weeks (eight baseline assessments).

**Figure 3:**
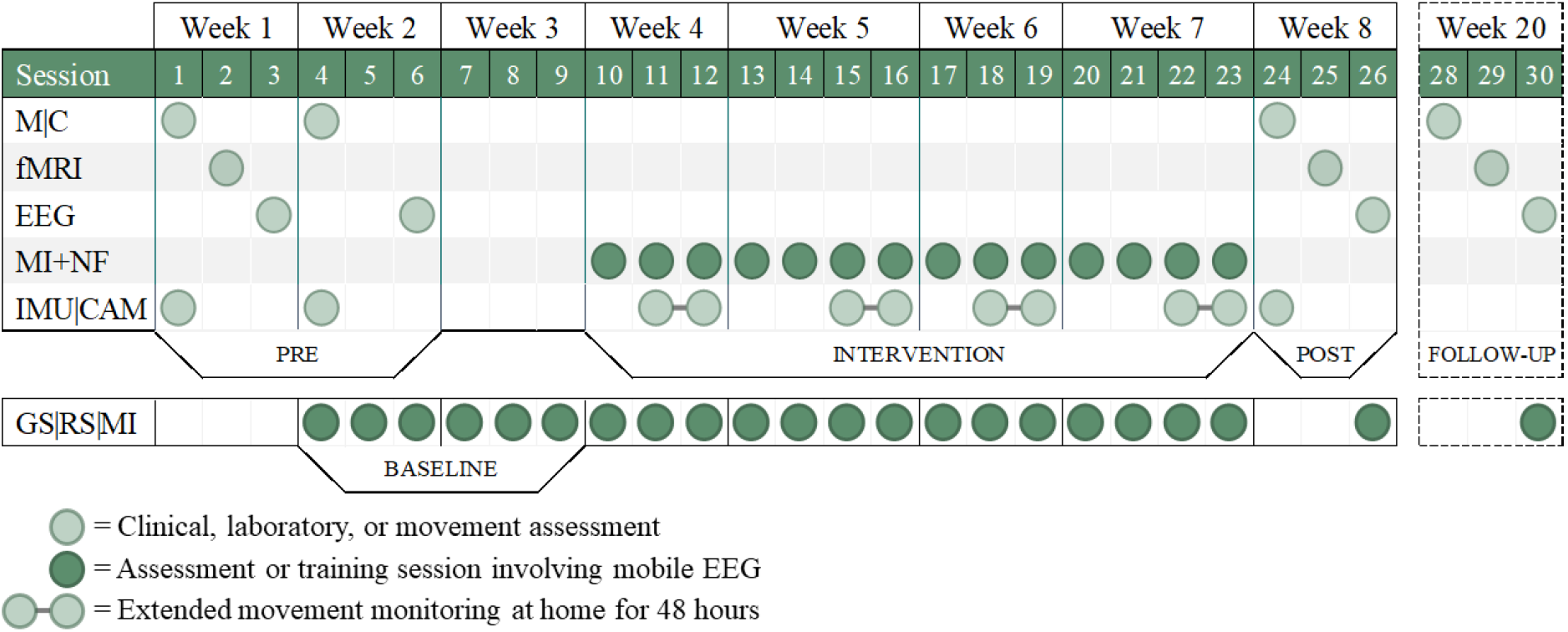
Study design. Overview of the study schedule and timing of all assessments and training sessions following a fixed two-day study rhythm. The timeline illustrates the pre-assessment phase, a variable-length baseline period, the four-week home-based intervention (MI+NF; motor imagery neurofeedback training), the post-assessment, and an exploratory three-month follow-up, shown here for a representative participant with six baseline sessions. The pre-, post, and follow-up assessments comprise measures of motor (M) and cognitive (C) function as well as functional magnetic resonance imaging (fMRI) and stationary electroencephalography (EEG). During the intervention phase, extended movement monitoring is conducted once per week, during which participants wear five inertial measurement units (IMU) continuously for 48 hours and record activities of daily living using a camera (CAM). Baseline measurements of grip strength (GS), resting-state EEG (RS), and motor imagery (MI) are repeated throughout the baseline and intervention phases. While the overall sequence of sessions is identical across participants, the total study duration varies depending on baseline length.

All study sessions take place from Monday to Sunday on a two-day cycle. This scheduling is particularly important for the intervention phase, which is designed as an intensive but well-paced form of brain training: participants train frequently, yet always have a rest day in between sessions to support memory consolidation (35, 36). Session times are arranged according to each participant’s preferences and daily routines. However, baseline and training sessions are conducted at the same time of day whenever possible, with exceptions limited to ±2 hours. This consistency is intended to maintain data quality and minimize time-of-day-related variability. The study is designed as an ecologically valid, home-based adjunct to participants’ everyday lives and their ongoing rehabilitation. Participants continue receiving their regular therapeutic care (e.g., physiotherapy, occupational therapy) without restrictions, as controlling or restricting usual care would undermine the naturalistic character of the intervention. Participants are explicitly instructed at enrolment to maintain their usual therapeutic routines unchanged throughout the entire study period – neither intensifying nor discontinuing ongoing care – in order to hold the rehabilitation context constant across baseline and intervention phases. Information on current health-related activities and treatments, including the type and frequency of ongoing therapeutic or rehabilitative care, is collected as part of the screening process and documented as contextual information in the study records, and reassessed after study completion to determine whether participants maintained their usual therapeutic routines during the study period.

Pre-, post-, and follow-up-assessments include comprehensive motor, cognitive, EEG, MRI, and movement measurements to capture behavioral and neural effects of training (see Sections 2.4.1 Motor and Cognitive Assessment; 2.4.2 Functional MRI; 2.4.4 Stationary High-Density EEG). To improve reliability, selected motor, cognitive, and stationary EEG assessments are conducted twice during the pre-assessment phase. The repetition is applied to measures where repeated testing is feasible within practical and participant-related constraints, in order to reduce the influence of day-to-day fluctuations related to factors such as fatigue, sleep quality, stress, or unfamiliarity with the testing environment. During the baseline phase, maximal grip strength of both hands, resting-state EEG, and a single run of MI without NF are recorded (see Section 2.4.3 MBD Baseline Measurement). These measurements continue throughout the entire intervention phase, where they are administered at the start of each session before the MI-NF training begins, and are repeated once more during the post-assessment, and, when applicable, during the Follow-Up. The intervention itself consists of intensive, mobile-EEG MI-NF sessions (MI-NF training) carried out three to four times per week over four weeks at home (see Section 2.4.5 Intervention: Motor Imagery Neurofeedback Training). In addition, extended movement monitoring using inertial measurement units (IMUs) and a camera is conducted once per week during the intervention phase to capture upper-limb movement behavior in daily life (see Section 2.4.6 Extended Movement Monitoring in Daily Life). The three-month follow-up assessment is exploratory in nature and is intended to investigate the persistence of potential training-related effects beyond the primary post-intervention assessment. As the follow-up serves an exploratory objective rather than a primary study objective, participation requires separate consent and is contingent upon participant willingness and logistical feasibility.

#### 2.3.1 Study Setting

All pre- and post-assessment sessions take place at the University of Oldenburg, Germany, within designated facilities including the EEG laboratory, Neuroimaging Unit, and behavioral testing rooms. The MI-NF training is delivered in participants’ homes across the region (primarily Oldenburg, Bremen, and surrounding areas). As this is a single-center study with home-based training components, the University of Oldenburg serves as the sole study site.

#### 2.3.2 Data Collection Methods

The MINTS study collects motor, cognitive, EEG, and MRI data using standardized procedures to ensure consistency and data quality. Motor and cognitive assessments include questionnaires and standardized tests. Scores are first recorded on paper and subsequently digitized using REDCap (Research Electronic Data Capture) tools hosted at the University of Oldenburg (37, 38). REDCap is a secure web-based platform that facilitates validated data entry, maintains audit trails, allows easy export to common statistical software, and supports integration with external data sources. For instruments administered repeatedly across the study (e.g., MoCA), alternate test versions are used to reduce learning effects and support validity. All verbal instructions, including those for MI-NF task performance, follow either fixed scripts or consistency in content to ensure consistent administration across participants. All assessors, including student research assistants, received in-person training from a licensed physiotherapist to ensure reliable administration and scoring of motor assessments. All MI-NF sessions are administered by trained study personnel during scheduled home visits and follow a standardized protocol. During these sessions, participants are behaviorally monitored, and relevant observations are documented. The training may be discontinued if participants decide to withdraw from the study, if persistent technical problems occur that cannot be resolved despite troubleshooting, or if external circumstances arise that prevent further participation, such as acute illness, hospitalization or comparable unforeseen events. If possible, participants who discontinue the training will still be invited to complete the post-assessment measures.

### 2.4 Experimental Procedure

#### 2.4.1 Motor and Cognitive Assessment

During the first study visit, several study-initiation procedures are completed before the motor and cognitive assessment begins. Participants provide written informed consent, undergo an aphasia screening, and complete an MRI safety questionnaire to confirm eligibility for MRI procedures. These procedures are followed by a brief overview of the study and an explanation of the session structure.

After these introductory steps, the motor and cognitive assessment starts. Participants first complete the 12-Item Short Form Health Survey (SF-12; Ware, Kosinski (39)), followed by the Montreal Cognitive Assessment (MoCA; Nasreddine, Phillips (40)), which is administered by the investigator. Subsequently, IMUs (Move 4, movisens GmbH, Karlsruhe, Germany) are attached to both upper and lower arms as well as the sternum to capture body movement. The IMU sensors record the three-dimensional acceleration and the angular rate at 64 Hz. The five IMUs are synchronized with each other before each session. Participants are informed that three cameras (Azure Kinect DK, Microsoft, Redmond, Washington, USA) will record their performance during the motor assessments: one positioned on the left side, one on the right side, and one mounted above the table. These cameras record color (resolution of 2048 x 1536) and depth (resolution of 320 x 288) information at 30 Hz. To synchronize IMUs, camera recordings, and task onset, participants are asked to sit still for five seconds, press a button, and then remain still for another five seconds before each motor assessment begins. The Modified Ashworth Scale (MAS; Bohannon and Smith (41)) is conducted, followed by the Fugl-Meyer Assessment for Upper Extremity (FMA-UE; Fugl-Meyer, Jaasko (42)). The 27-item structure used in this study excludes the three reflex activity items (biceps/finger flexor reflexes and triceps reflex) and the three coordination/speed items (tremor, dysmetria, and time). It has demonstrated good structural validity and internal consistency (43). Afterwards, participants are offered a short break (5-10 minutes) if desired. Testing then continues with the Action Research Arm Test (ARAT; Lyle (44)), the Jebsen-Taylor Hand Function Test (JHFT; Jebsen, Taylor (45)), the Modified Frenchay Scale (MFS; based on the Frenchay Arm Test by Wade, Langton-Hewer (46); modified version described in Gracies, Hefter (47)), and the Test of Attentional Performance - Mobility Version (TAP-M; Zimmermann and Fimm (48)), including the subtests Distractibility, Executive Control, Divided Attention, and Go/NoGo. A second optional break (5-10 minutes) is offered at this point. The session concludes with the Functional Gait Assessment (FGA; Wrisley, Marchetti (49)), the Stroke Impact Scale - Version 3.0 (SIS; Carod-Artal, Coral (50)), and the Beck Depression Inventory-II (BDI-II; Beck, Steer (51)). All IMUs are removed at the end of testing. The order of assessments was chosen to distribute motor and cognitive demands across the session and to minimize fatigue. Short breaks were offered at predefined points to maintain participant comfort and data quality.

The motor and cognitive assessment is conducted twice prior to the start of the intervention. Pre-I comprises the full test battery, while Pre-II includes a reduced set of assessments. Tests that are not expected to show meaningful change over short time intervals, or for which repeated administration within a short amount of time is not appropriate, are omitted in Pre-II. After completion of the intervention period, all assessments are repeated the subsequent week (Post). Participants who indicated in the informed consent at study entry that they were willing to be contacted for a follow-up assessment are invited to complete an additional assessment three months later (Follow-Up), with appointments scheduled after completion of the last study session. A detailed overview of which tests are conducted in each session is provided in Table 3.

**Table 3:** Overview of motor and cognitive tests and questionnaires across assessment time points.

|  |  | Pre-I | Pre-II | Post | Follow-Up |
| --- | --- | --- | --- | --- | --- |
| **12-Item Short Form Health Survey:** Self-report questionnaire assessing general physical and mental health-related quality of life | SF-12 | X |  | X | X |
| **Montreal Cognitive Assessment:** Screening tool for global cognitive functioning across different domains | MoCA | X | X | X | X |
| **Modified Ashworth Scale:** Clinical measure of muscle tone and spasticity in the upper limb | MAS | X | X | X | X |
| **Fugl-Meyer Assessment for Upper Extremities:** Standardized evaluation of motor impairment | FMA-UE | X | X | X | X |
| **Action Research Arm Test:** Performance-based assessment of upper-limb function on grasp, grip, pinch, and gross movement | ARAT | X | X | X | X |
| **Jebsen-Taylor Hand Function Test:** Timed assessment of everyday manual tasks evaluating fine and gross manual dexterity | JHFT | X | X | X | X |
| **Modified Frenchay Scale:** Structured rating of the frequency and quality of affected-arm use in daily life | MFS | X | X | X | X |
| **Test battery for Attention Assessment Mobility Version:** Computerized assessment of attentional capacities | TAP-M | X |  | X | X |
| **Functional Gait Assessment:** Clinical evaluation of walking ability under complex gait conditions | FGA | X |  | X | X |
| **Stroke Impact Scale – Version 3.0:** Patient-reported outcome measure assessing perceived stroke-related disability | SIS | X |  | X | X |
| **Becks Depression Inventory-II:** Validated questionnaire measuring depressive symptoms | BDI-II | X |  | X | X |

#### 2.4.2 Functional MRI

MRI data are acquired at up to three time points across the study: during the pre-assessment phase (Pre), after completion of the intervention (Post), and, where applicable, during an exploratory three-month follow-up assessment (Follow-Up). Each MRI session follows an identical acquisition protocol consisting of five segments (fMRI task, T1, DWI, resting-state, FLAIR). Prior to the MRI session, participants provide written confirmation of MRI eligibility and informed consent. They are informed about the general structure of the MRI session, including the acquisition of multiple imaging protocols and the performance of a motor execution/attempt task and a resting-state measurement. Given the total scan duration of approximately one hour, the MRI session is divided into two parts with a scheduled break of 10–15 minutes to minimize fatigue-related movement effects.

MRI data are acquired on a 3 Tesla whole-body Siemens Magnetom Prisma scanner equipped with a 64-channel head coil. Data acquisition starts with the fMRI motor execution/attempt task. During this task, participants are visually prompted by a graphic (blue areas either on the left or right side) presented for five seconds on a screen via a mirror system. Participants are instructed to repeatedly open and close the indicated hand, or attempt to do so, for the duration of the prompt. Task prompts target both the affected and unaffected hand with equal probability. The motor execution/attempt task lasts approximately 10 minutes and comprises a total of 75 trials, including 25 left-hand, 25 right-hand, and 25 rest trials in which a central point is shown and both hands are to be kept motionless (see Figure 4). Trial order is pseudorandomized and was defined prior to the study, with the constraint that no trial type occurs more than twice consecutively. This constraint was applied to prevent fatigue or strain of the affected hand due to chance trial sequences, which could otherwise influence task performance and neural responses. The same predefined sequence is used for all participants.

**Figure 4:**
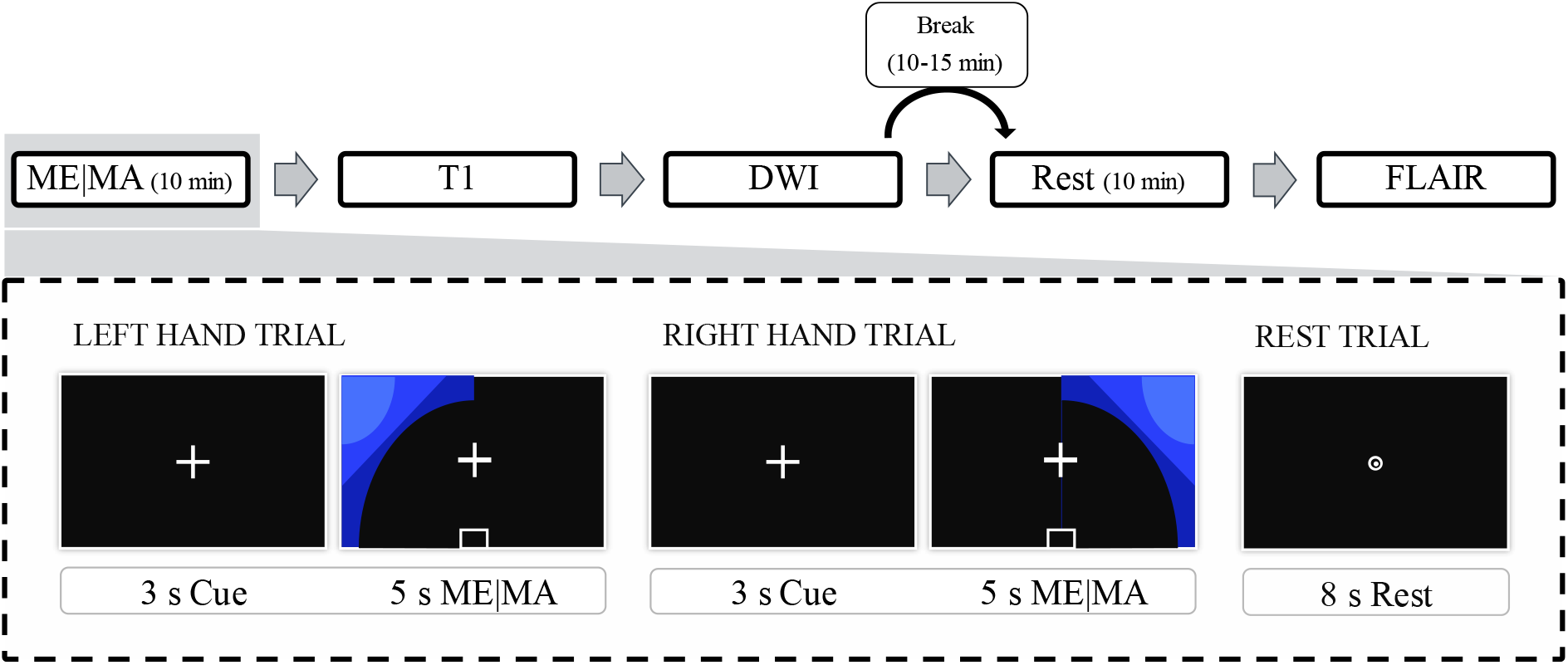
MRI sessions including motor execution/attempt task and imaging sequence order. Schematic overview of the MRI session, with the upper part illustrating the sequence of acquisitions and the lower part detailing the motor execution/attempt (ME/MA) task. The session begins with a ME/MA task, followed by structural T1-weighted imaging (T1) and diffusion weighted imaging (DWI). After DWI acquisition, a scheduled break of 10-15 minutes is provided before the resting-state fMRI measurement (Rest). The session concludes with a fluid-attenuated inversion recovery (FLAIR) sequence. The ME/MA task consists of left-hand, right-hand, and rest trials. Each motor trial includes a 3-s visual cue followed by a 5-s ME/MA period, during which participants repeatedly open and close, or attempt to open and close, the prompted hand. Rest trials consist of an 8-s rest period in which both hands are to be kept motionless. Trials are presented in a pseudorandomized order.

Following the fMRI task, a high-resolution T1-weighted structural image and diffusion-weighted imaging (DWI) are acquired. After the scheduled break, resting-state fMRI data are collected for 10 minutes. Participants are instructed to keep their eyes open, fixate a central point, remain still, stay awake, and relax without engaging in any specific task. Finally, a fluid-attenuated inversion recovery (FLAIR) sequence is acquired, after which the session is concluded.

Structural images are acquired using a 3D T1-weighted sequence (TR = 2000 ms, TE = 2.07 ms, flip angle = 9, slice thickness = 0.8 mm, voxel size = 0.8 x 0.8 x 0.8 mm^3^, 224 slices, field of view = 240 x 240 mm). Functional MRI data for the motor execution/attempt task and resting-state are acquired using a 2D multiband echo-planar imaging (EPI) sequence with blood oxygen level-dependent (BOLD) contrast, from the Center for Magnetic Resonance Research (CMRR), University of Minnesota (52–54) (TR = 850 ms, TE = 30 ms, flip angle = 62, distance factor = 0%, slice thickness = 2.5 mm, field of view = 192 x 192 mm, matrix = 76 x 76, voxel size = 2.5 x 2.5 x 2.5 mm³, 48 slices, multiband factor = 4, volumes for motor execution/attempt task = 1052, volumes for resting-state = 750). Diffusion-weighted images are acquired with 12 non-diffusion-weighted images (b = 0 s/mm²) and 116 diffusion-weighted images (b = 2000 s/mm²) (TR = 3428 ms, TE = 79 ms, flip angle = 76, slice thickness = 1.5 mm, field of view = 192 x 192 mm, voxel size = 1.5 x 1.5 x 1.5 mm^3^, 75 slices, 116 directions, multiband factor = 3). A FLAIR sequence was acquired to characterize lesion location and extent (TR = 5000 ms, TE = 390 ms, TI = 1800 ms, slice thickness = 1.0 mm, voxel size = 1.0 × 1.0 × 1.0 mm³, field of view = 256 × 256 mm, 160 slices).

#### 2.4.3 MBD Baseline Measurements

MBD baseline measurements include assessments of motor performance and neural activity, comprising maximal hand grip strength as well as resting-state and task-related EEG recordings. These measures were selected because they can be assessed repeatedly across baseline and intervention phases with limited participant burden while capturing behavioral and neurophysiological domains relevant to the intervention. Resting-state EEG provides a measure of functional connectivity independent of task performance, whereas MI without NF allows assessment of MI-related sensorimotor activity without and immediately before concurrent NF. Grip strength serves as a brief and objective indicator of motor performance. It is not intended to represent upper-limb motor function in its entirety, but complements the broader clinical and movement-based motor assessments collected before and after the intervention. Baseline measurements start in parallel with the pre-assessment phase of the study. Specifically, baseline data acquisition begins from the second pre-assessment week onward, resulting in a temporal overlap between pre-assessment and baseline measurements. Following neuroradiological classification, participants are assigned to one of the predefined lesion-location groups. The assignment also specifies the number of baseline sessions for a given participant (see Section 2.2.4 Randomization). Consequently, the duration of the baseline phase and thus the overall study duration differs across participants in accordance with the implemented MBD.

Maximal grip strength is assessed using a Baseline® BIMS Digital Grip Dynamometer (clinical model; Fabrication Enterprises Inc., White Plains, New York, USA). Grip strength is measured six times in total (three trials per hand), starting with the less affected hand. Short breaks are provided between trials. Participants hold the dynamometer with the arm extended and are instructed to squeeze as forcefully as possible. All values are recorded, and mean grip strength is calculated separately for each hand for subsequent analyses.

EEG data are recorded using a mobile amplifier (SMARTING mobi, mBrainTrain, Belgrade, Serbia) in combination with a 24-channel mobile EEG cap with a motor-area layout (EasyCap, Herrsching, Germany). Data are sampled at 500 Hz, and electrode impedances are maintained below 1 0 kΩ. Resting-state EEG is recorded for 10 minutes using SMARTING Streamer software (version 3.4.3; mBrainTrain, Belgrade, Serbia). Participants are instructed to sit still, fixate a central point on the screen, and remain relaxed without engaging in any specific cognitive task. Following the resting-state recording, the amplifier is connected via Bluetooth to OpenViBE Server (version 0.17.1; Inria, Rennes, France; Renard, Lotte (55)). Then, participants perform an MI run without NF. Participants are instructed to repeatedly imagine opening and closing their hand in a kinesthetic manner, focusing on the sensory experience associated with the movement. To facilitate vivid MI, participants briefly physically perform the opening and closing movement with their less affected hand and attempt it with the affected hand prior to the MI task. The MI run lasts approximately 10 minutes and consists of 40 trials (20 left-hand and 20 right-hand trials) presented in pseudorandomized order. Each trial begins with a 5-s baseline period, followed by a 3-s preparation cue. During the subsequent 5-s MI interval, a visual prompt is presented on either the left or right side of the screen, indicating the hand to be imagined. Trials are separated by a variable inter-trial interval ranging from 0 to 4 s (see Figure 5A).

**Figure 5:**
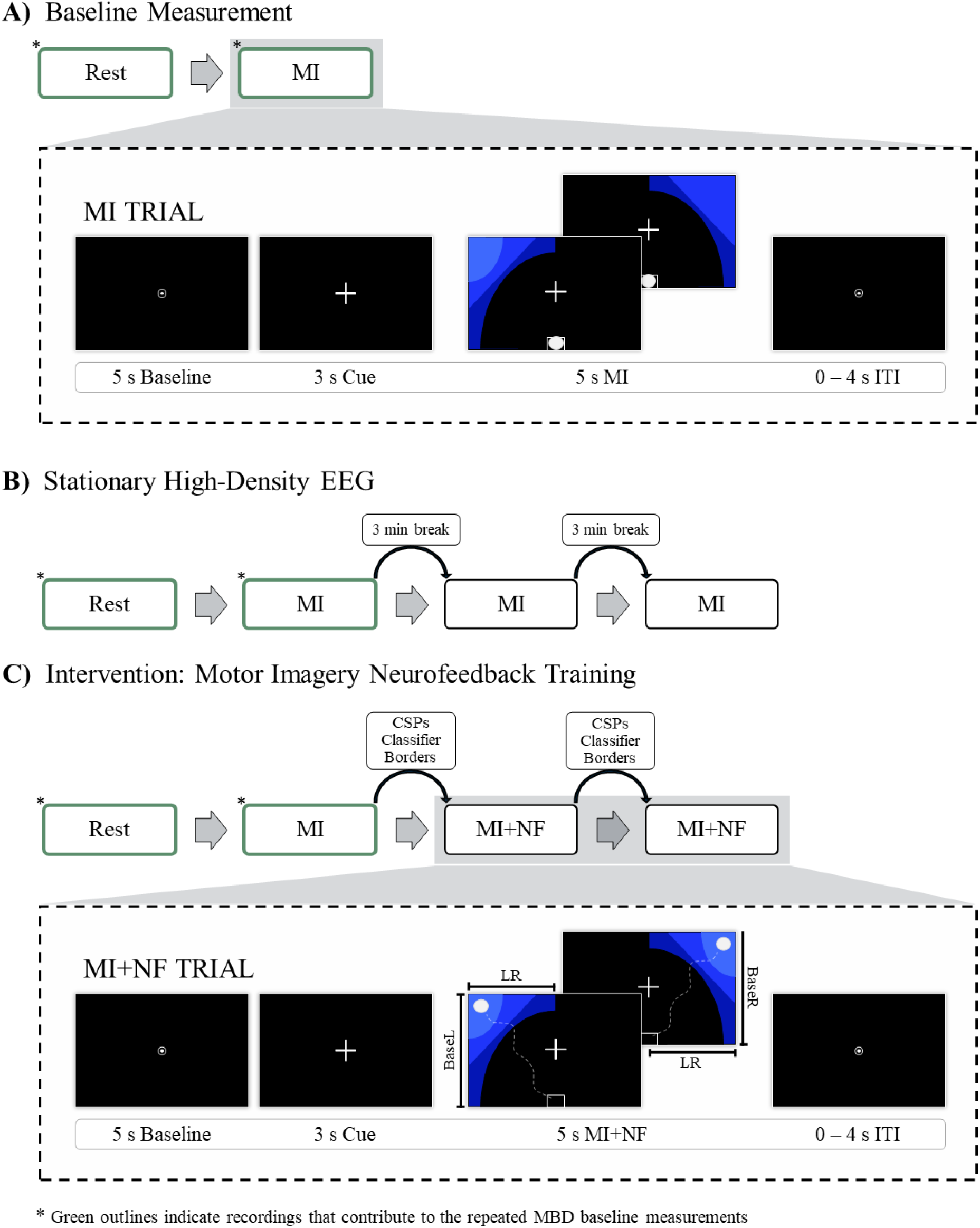
EEG sessions including baseline measurements, stationary EEG, and motor imagery neurofeedback training procedure. **(A)** Each session begins with a resting-state (Rest) recording followed by one motor imagery (MI) run. Each MI trial consists of a 5-s baseline period, a 3-s preparation cue, a 5-s MI interval, and a variable inter-trial interval (ITI). **(B)** After the resting-state recording, participants perform three consecutive MI runs following the trial structure shown in panel A. **(C)** Following the MBD baseline measurement (resting-state and MI run without neurofeedback), two MI runs with neurofeedback (MI+NF) are performed. EEG data from the MI run are used to compute common spatial patterns (CSPs) and train participant-specific classifiers (BaseL, BaseR, LR) and borders. These parameters control real-time feedback during MI+NF trials and are recalculated after the first neurofeedback run for the subsequent run. The MI+NF trial follows the same temporal structure as the MI trial but includes continuous visual feedback reflecting task-related sensorimotor activity (adapted from Decker, Daeglau (60)).

#### 2.4.4 Stationary High-Density EEG

Stationary high-density EEG recordings are used to assess neural activity under controlled laboratory conditions at three or four time points during the study: twice prior to the intervention (Pre-I and Pre-II), after completion of the intervention (Post), and, where applicable, during Follow-Up, mirroring the schedule of the motor and cognitive assessments. The overall layout of these sessions is identical with a few exceptions: During the first stationary EEG session (Pre-I), prior to EEG preparations, participants are introduced to the experimental setup and the overall procedure. Also, only during Pre-I, The Kinesthetic and Visual Imagery Questionnaire (KVIQ) is administered (56). The KVIQ provides a subjective measure of MI ability and captures participants’ self-evaluations of visual and kinesthetic imagery experiences. At the Post session, following the EEG measurement, participants complete a study completion questionnaire assessing practical implementation, acceptability, participant burden, usability of the home-based setup, perceived training effects, and overall satisfaction with the intervention.

EEG data are recorded using a BrainAmp amplifier system (Brain Products GmbH, Gilching, Germany) with a 64-channel EEG cap (EasyCap, Herrsching, Germany) arranged in an equidistant layout. A nose-tip electrode is used as reference, and a frontopolar electrode serves as ground. Two electrodes placed below the eyes record eye movements and blinks. Signals are sampled at 500 Hz, and electrode impedances are kept below 1 0 kΩ. Surface electromyography (EMG) is recorded bilaterally to monitor muscle activity during MI. EMG electrodes are placed over hand and forearm muscles, including the first dorsal interosseous muscle, the flexor digitorum superficialis, and the abductor pollicis longus, with reference and ground electrodes positioned at the clavicles. EMG impedances are maintained below 1 00 kΩ. The EMG recordings are used as a control measure to monitor overt or unintended muscle activity during MI, as participants are instructed to perform MI without actual movement. Depending on the observed data quality and activity patterns, EMG information may be used for trial-level quality control, including the potential exclusion of affected MI trials from EEG analyses, as well as for sensitivity analyses or to support the interpretation of participant-specific activity patterns during MI.

Prior to data acquisition, the MI task is explained in detail. Participants are instructed to repeatedly imagine opening and closing their hand in a kinesthetic manner, with an emphasis on the tactile sensations associated with the movement, rather than the visual image. To facilitate vivid MI, participants briefly physically perform the opening and closing movement with the less affected hand and attempt the movement with the affected hand before the EEG recording begins. Each session begins with a 5-minute resting-state EEG recording. Participants are instructed to sit still, fixate a central point on the screen, and remain relaxed without engaging in any specific cognitive task. Resting-state data are acquired using BrainVision Recorder software (version 1.20.0506; Brain Products GmbH, Gilching, Germany). Subsequently, MI data are recorded using OpenViBE Server (version 0.17.1; Inria, Rennes, France). Participants complete three runs of MI without additional NF. Rest periods of approximately three minutes are provided between runs to minimize fatigue. Each run lasts approximately 10 minutes and follows the same trial structure as described for the baseline MI measurements (see Figure 5B). During the Pre-II, Post, and, when applicable, Follow-Up assessments, the stationary EEG session is combined with the MBD baseline measurement to reduce participant burden. In these sessions, the resting-state recording is extended to 10 minutes, allowing the first 5 minutes to be used for the stationary EEG analyses and the full recording to contribute to the MBD resting-state EEG outcome. Similarly, the first MI run is used both as part of the stationary EEG assessment and as the corresponding MBD task-EEG measurement.

#### 2.4.5 Intervention: Motor Imagery Neurofeedback Training

Each intervention session follows the same structure as the MBD baseline measurements described in Section 2.4.3, beginning with assessment of maximal hand grip strength, EEG preparation (including reduction of electrode impedances to below 1 0kΩ), resting-state EEG, and one MI run without NF, before proceeding to two additional MI runs with real-time NF as the training component. This initial MI run serves as a calibration run for the subsequent NF. Following the initial MI run, two MI runs with real-time NF are performed as training (see Figure 5C). The NF is provided as a two-dimensional visual display consisting of a ball presented in the lower center of the screen. Participants are instructed to control the ball using kinesthetic MI of repeated hand opening and closing, with the aim to guide it toward the upper left or upper right corner of the screen, into the center of the corresponding blue target area. NF is based on imagery-related modulation of sensorimotor activity during kinesthetic MI and provides continuous information about task-related brain activity.

EEG data recorded during the initial MI run without NF are used to calibrate the NF parameters. Raw EEG signals from all 24 channels of the mobile EEG cap are band-pass filtered in the mu and beta (8–30 Hz) frequency ranges, which are known to reflect sensorimotor rhythm modulation during MI. Data are segmented into epochs ranging from 0.5 to 4.5 s after MI onset and labeled according to left- and right-hand MI. Epochs containing excessive artifacts are excluded from further processing using a joint probability-based rejection criterion (EEGLAB function pop_jointprob, threshold = 3 standard deviations).

Common Spatial Patterns (CSP) analysis is performed in MATLAB (version R2022b; MathWorks, Natick, Massachusetts, USA, RID:SCR_001622) using the EEGLAB toolbox (version 14.1.1.) (57) with a CSP pipeline (58) to derive spatial filters that optimally discriminate between left- and right-hand MI. CSP-based feature extraction is among the most commonly applied approaches in EEG-based MI-BCI research using wearable technologies (59) and has been applied in an MI-NF paradigm in previous studies from our group (24, 60–63). CSP is used to enhance task-relevant sensorimotor features by maximizing variance differences between the two imagery conditions while attenuating common background activity. For each calibration run, the most neurophysiologically plausible CSP components (one reflecting left-hand and one reflecting right-hand MI) are selected based on their spatial distribution over sensorimotor areas. The resulting CSP filter coefficients are then transferred to OpenViBE (version 0.17.1) for online processing. Within OpenViBE, EEG signals are spatially filtered using the CSP coefficients and temporally filtered using a band-pass filter between 8 and 30 Hz. For classifier training, MI-related activity is extracted from epochs between 0.5 and 4.5 s after cue onset, while baseline activity is derived from pre-cue intervals (5 s baseline) preceding MI onset. Features are computed as logarithmic band power values calculated in overlapping time windows and are used to train linear discriminant analysis (64) classifiers with seven-fold cross-validation. Three classifiers control the movement of the NF ball. Horizontal movement reflects hemispheric lateralization and is based on the difference between left- and right-hand MI activity (classifier LR). Vertical movement is governed by two classifiers, which quantify the difference between task-related sensorimotor activity during MI and baseline activity for left-hand (classifier BaseL) and right-hand (classifier BaseR) imagery, respectively. Border values defining the dynamic range of the NF display are computed from classifier outputs and are used to scale the visual feedback for each participant. After completion of the first NF run, EEG data from this run are used to update CSP filters, classifiers, and border values. These updated parameters are then applied to the second NF run, allowing for adaptive recalibration to account for intra-session changes in neural activation patterns and to optimize NF performance.

#### 2.4.6 Extended Movement Monitoring in Daily Life

At four predefined time points during the intervention phase, participants undergo extended movement monitoring in their daily environment. Immediately following the respective MI-NF training session, five IMUs are attached to the upper body and upper limbs, using the same sensors and configuration as during the standardized motor and cognitive assessments (see Section 2.4.1 Motor and Cognitive Assessment). Participants wear the IMUs for approximately 48 hours to capture spontaneous, task-independent upper-limb movements during everyday activities outside the training context. Participants may remove the sensors temporarily for showering or sleeping if desired. The sensors are removed two days later, immediately before the subsequent MI-NF training session, ensuring uninterrupted movement recording while maintaining consistency with the intervention schedule.

In parallel, participants are provided with a camera (GoPro Hero11 Black, GoPro, Inc., San Mateo, California, USA) for the same monitoring period. The camera is placed in the kitchen and records color video at a resolution of 1920 × 1080 pixels and 30 Hz. Participants are instructed to independently record short video sequences while performing routine activities of daily living, with a particular focus on kitchen-related tasks (e.g., food preparation). Video recordings are entirely participant-controlled. Participants independently decide when recordings are started and stopped, and only activities they explicitly choose to share are captured. No continuous video monitoring is performed. Participants may decline or limit recordings at any time. The camera recordings are intended to capture functional upper-limb use in a naturalistic setting and complement the IMU-based kinematic data.

The combined IMU and video recordings allow assessment of real-world movement behavior beyond structured test situations. This approach enables exploratory analyses of whether changes in everyday upper-limb use can be detected over the course of the intervention and whether these changes correspond to those observed in clinical assessments and laboratory-based measures.

### 2.5 Outcomes

The outcomes of this study are designed to capture training-related changes in neural activity, functional brain connectivity, and motor performance following MI-NF training. Outcomes are structured into primary, secondary, tertiary, and exploratory, integrating neurophysiological, neuroimaging, and behavioral assessments to comprehensively characterize intervention effects. An overview of all outcome measures is provided in Table 4 and the following sections describe the individual outcome domains in detail.

**Table 4:**
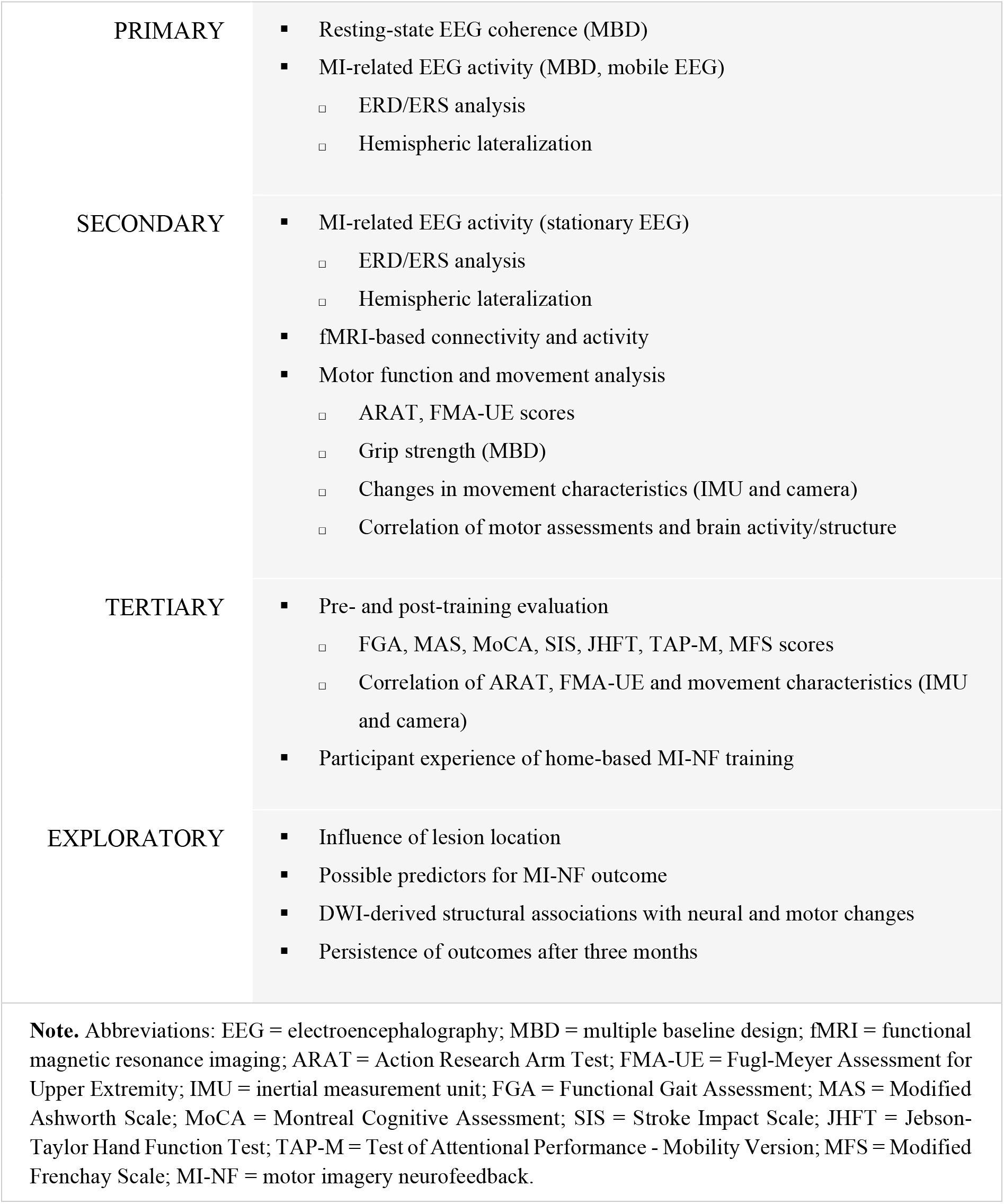
Study outcomes.

|  |  |
| --- | --- |
| PRIMARY | ▪ Resting-state EEG coherence (MBD)<br>▪ MI-related EEG activity (MBD, mobile EEG)<br>▫ ERD/ERS analysis<br>▫ Hemispheric lateralization |
| SECONDARY | ▪ MI-related EEG activity (stationary EEG)<br>▫ ERD/ERS analysis<br>▫ Hemispheric lateralization<br>▪ fMRI-based connectivity and activity<br>▪ Motor function and movement analysis<br>▫ ARAT, FMA-UE scores<br>▫ Grip strength (MBD)<br>▫ Changes in movement characteristics (IMU and camera)<br>▫ Correlation of motor assessments and brain activity/structure |
| TERTIARY | ▪ Pre- and post-training evaluation<br>▫ FGA, MAS, MoCA, SIS, JHFT, TAP-M, MFS scores<br>▫ Correlation of ARAT, FMA-UE and movement characteristics (IMU and camera)<br>▪ Participant experience of home-based MI-NF training |
| EXPLORATORY | ▪ Influence of lesion location<br>▪ Possible predictors for MI-NF outcome<br>▪ DWI-derived structural associations with neural and motor changes<br>▪ Persistence of outcomes after three months |
| **Note.** Abbreviations: EEG = electroencephalography; MBD = multiple baseline design; fMRI = functional magnetic resonance imaging; ARAT = Action Research Arm Test; FMA-UE = Fugl-Meyer Assessment for Upper Extremity; IMU = inertial measurement unit; FGA = Functional Gait Assessment; MAS = Modified Ashworth Scale; MoCA = Montreal Cognitive Assessment; SIS = Stroke Impact Scale; JHFT = Jebson-Taylor Hand Function Test; TAP-M = Test of Attentional Performance - Mobility Version; MFS = Modified Frenchay Scale; MI-NF = motor imagery neurofeedback. |  |

#### 2.5.1 Resting-State EEG Coherence

Resting-state EEG coherence is one of the primary outcome measures of the study. EEG coherence analysis is used to quantify functional connectivity. It is a frequency-domain method that estimates the degree of synchronization between pairs of EEG signals. Coherence values range from 0 (no synchronization) to 1 (high synchronization), reflecting the strength of coupling between brain regions (65). In the present study, resting-state EEG coherence will be quantified primarily in motor-relevant frequency bands, with a focus on the alpha/mu (8–13 Hz) and beta (13–30 Hz) frequency ranges. Analyses will primarily focus on connectivity patterns involving motor and premotor regions as well as prefrontal regions, including ipsilesional and contralesional sensorimotor areas and prefrontal electrode clusters. In addition to these predefined regions and frequency bands of interest, exploratory data-driven coherence analyses may be conducted across available electrode pairs and frequency ranges to characterize broader training-related network changes.

Resting-state EEG is assessed repeatedly within the MBD, with 3 to 8 measurements during the baseline phase and 14 measurements during the intervention phase. The primary analysis therefore focuses on temporal changes in resting-state EEG coherence after intervention onset compared with the preceding baseline phase, rather than on post-intervention values alone. Training-related increases or decreases in coherence will be interpreted as modulation of neural networks potentially involved in MI and NF learning.

#### 2.5.2 MI-Related EEG Activity (MBD, Mobile EEG)

MI-related EEG activity derived from mobile EEG recordings is a primary outcome of the study and is assessed repeatedly within the MBD across the baseline and intervention phases. The primary analysis focuses on whether MI-related EEG activity changes systematically after intervention onset compared with the preceding baseline phase.

MI-related EEG activity is quantified using two related but distinct EEG-derived measures: EEG power, operationalized as ERD/ERS, and hemispheric lateralization. ERD/ERS reflects motor-related oscillatory activity in the mu and beta frequency bands (8–30 Hz) and will be reported in µV² or dB (66). ERD/ERS values will be calculated relative to a predefined within-trial reference interval, thereby accounting for inter-individual differences in absolute EEG power and enabling comparison across sessions and participants. Hemispheric lateralization will be calculated from ERD/ERS values as the difference in power between the ipsilesional and contralesional hemispheres.

Within the MBD, task-related mobile EEG during a single run of MI without NF is assessed repeatedly across 3 to 8 baseline measurements and 14 intervention sessions. This allows the examination of temporal changes in MI-related ERD/ERS and hemispheric lateralization across the intervention period. Training-related changes in these measures following intervention onset are interpreted as modulation of motor-related cortical activation patterns and provide insight into how MI-NF training affects neural activity over time.

#### 2.5.3 MI-Related EEG Activity (Stationary EEG)

MI-related EEG activity is additionally assessed using stationary high-density EEG recordings. These recordings are not part of the MBD but serve as secondary outcomes for pre- and post-intervention comparisons and, where available, exploratory follow-up analyses. They are used to further characterize training-related changes in hemispheric motor-related activity independent of NF under more controlled laboratory conditions.

During the stationary EEG assessments, participants perform three MI runs without NF. These runs are recorded during two pre-assessment measurements, one post-intervention measurement, and an additional measurement during the exploratory three-month follow-up assessment. These stationary EEG recordings are not part of the MBD but serve as secondary outcomes for pre- and post-intervention comparisons. Consistent with the mobile EEG-based analyses, stationary EEG data will be used to examine MI-related ERD/ERS and hemispheric lateralization during MI without NF, thereby further characterizing changes in hemispheric motor-related activity independent of NF. Literature suggests that following a stroke, ipsilesional, contralateral motor-related activity is often reduced compared to contralesional ipsilateral regions (67–69). In this context, increases in ipsilesional ERD and shifts toward more balanced or ipsilesionally driven lateralization are interpreted as potentially beneficial adaptations associated with motor recovery (20, 22, 24).

#### 2.5.4 fMRI-Based Connectivity and Activity

fMRI-based connectivity and activity are secondary outcomes of the study. These measures are derived from BOLD signals acquired before the intervention (Pre), after the intervention (Post), and, where applicable, during the exploratory three-month follow-up assessment (Follow-Up). fMRI-based activity is assessed as regional BOLD signal changes during task performance, while functional connectivity is computed based on temporal correlations in BOLD signal fluctuations. Regions of interest will include motor and prefrontal areas (e.g., ipsilesional and contralesional motor cortex and dorsolateral prefrontal cortex). Connectivity analyses result in matrices representing large-scale brain network organization. Changes in fMRI-based activity and connectivity between pre- and post-intervention measurements are analyzed to identify training-related alterations in regional activation and network-level organization, providing complementary information to EEG-based outcomes.

#### 2.5.5 Motor Function and Movement Analysis

Motor function and movement analysis are secondary outcomes of the study and include clinical motor assessments, grip strength, and quantitative movement characteristics. Clinical motor outcomes are assessed using the ARAT and the FMA-UE at up to four time points (Pre-I, Pre-II, Post, and, where applicable, Follow-Up). Both assessments are widely used and well-established measures of upper-limb recovery after stroke (70). Although they are strongly related and have been shown to correlate highly with one another (71, 72), they differ in their assessment approach, scoring structure, and potentially their responsiveness to change in different patient populations and intervention settings (73). Given the expected heterogeneity of the MINTS study population, including a broad range of impairment severities and recovery profiles, one assessment may capture changes or limitations that are less apparent in the other. The combined use of both measures therefore allows a more comprehensive characterization of upper-limb motor recovery across participants. In addition, including both assessments facilitates comparison with previous stroke rehabilitation studies, which commonly report either ARAT or FMA-UE outcomes (70), and supports harmonization with future studies using comparable clinical motor assessment batteries. Accordingly, improvements in ARAT and FMA-UE scores are interpreted as improvements in upper-limb motor recovery.

Grip strength is assessed as part of the MBD, with 3 to 8 baseline measurements and 14 intervention sessions, and is calculated as the arithmetic mean of three trials per side. This repeated assessment allows the examination of temporal changes in motor performance after intervention onset compared with the preceding baseline phase.

Quantitative movement characteristics are derived from IMUs and camera-based recordings, from which skeleton data are extracted. These measurements are obtained at the same time points as the clinical motor assessments, with additional IMU recordings acquired four times during the intervention period (once per week). Movement parameters are derived from IMU and skeleton data, including acceleration, angular velocity, and joint position data from the upper limbs, with a focus on the affected upper limb. Derived measures may include indicators of movement amount, range of motion, movement velocity, and interlimb differences between the affected and less affected upper limb. For real-world recordings acquired at home, within-participant comparisons between the affected and less affected upper limb will be explored, as interlimb differences may provide information about movement behavior that is less dependent on the specific activities performed. Where appropriate, movement parameters will be normalized within participants, for example relative to the less affected upper limb or to individual baseline values, to improve comparability across sessions and account for inter-individual differences in movement capacity. Changes in these measures are interpreted as alterations in movement quality and motor control, reflecting potential improvements in coordination and range of motion.

In addition, correlations between motor assessments (ARAT, FMA-UE, grip strength) and measures of brain activity and structure are examined across pre- and post-intervention time points. These analyses are used to assess the relationship between neural changes and functional improvements, providing insight into whether observed neurophysiological alterations are associated with clinically meaningful motor recovery.

Together, these measures provide clinically relevant indicators of upper-limb motor function and allow evaluation of whether MI-NF training translates into meaningful functional improvements, reflected by higher clinical scores, increased grip strength, and enhanced movement range.

#### 2.5.6 Pre- and Post-Training Evaluation

Pre- and post-training evaluations of motor and cognitive functions are tertiary outcomes of the study. These include score-based measures (FGA, MAS, MoCA, SIS, MFS) and time-based measures (JHFT, TAP-M), which are assessed before and after the intervention. These measures are used to examine whether MI-NF training is associated with broader improvements in motor and cognitive performance beyond the primary and secondary outcomes. Improvements are reflected by increased scores and reduced completion times, indicating enhanced motor function, mobility, or cognitive performance.

Additionally, correlations between clinical motor outcomes (ARAT, FMA-UE) and quantitative movement characteristics derived from IMU and camera recordings are analyzed based on pre- and post-intervention measurements. These analyses are used to assess the 42 relationship between standardized clinical performance and objective movement characteristics. Consistent changes across these modalities are interpreted as indicators of a coherent and clinically meaningful response to MI-NF training.

#### 2.5.7 Participant Experience of Home-Based MI-NF Training

Participant experience is a tertiary outcome of the study. It is assessed after completion of the intervention using a study completion questionnaire, complemented by descriptive study-related indicators including participant retention, adherence to the planned training schedule, and completion of assessments and training sessions. The questionnaire addresses acceptability, integration of the training into daily life, usability of the home-based setup, participant burden, satisfaction with study procedures, perceived benefits, and willingness to continue or recommend the training. Together, these measures are used to describe participants’ experience with intensive home-based MI-NF training and its integration into everyday life. High participant retention, successful completion of planned assessments and training sessions, and predominantly positive participant ratings are interpreted as indicators that the intervention was acceptable to the target population and compatible with participants’ everyday routines.

#### 2.5.8 Exploratory Directions

Given the multimodal and longitudinal nature of the MINTS study, the dataset will allow additional exploratory analyses beyond the predefined analyses described above. The combination of neuroimaging, electrophysiological, behavioral, and real-world movement data may enable further examination of factors related to variability in neural and behavioral changes observed across participants. Potential exploratory work may include examining associations between lesion location (cortical, subcortical, or combined cortical/subcortical) and observed patterns in neural or behavioral measures. In addition, DWI-derived measures of white matter integrity and structural connectivity in motor-relevant pathways, particularly the corticospinal tract, may be explored in relation to changes in brain activation, functional connectivity, MI-related EEG activity, and clinical motor outcomes. Further exploratory analyses may also examine the persistence of observed changes at the exploratory three-month follow-up.

### 2.6 Data Analysis

Data analysis is structured according to the hierarchical outcome framework of the study and is aligned with the objectives described in Section 1.2. The first primary objective, which addresses training-related changes in resting-state functional connectivity, will be evaluated using resting-state EEG coherence derived from repeated mobile EEG measurements within the MBD. The second primary objective, which addresses MI-related brain activity independent of NF, will be evaluated using MI-related ERD/ERS and hemispheric lateralization derived from repeated mobile EEG recordings during MI without NF within the MBD. For both primary objectives, analyses will focus on whether changes in the respective dependent variables occur systematically after intervention onset compared with the preceding baseline phase. Secondary objectives will be evaluated using pre- and post-intervention comparisons of stationary EEG, fMRI, clinical motor assessments, and quantitative movement measures. These analyses are intended to further characterize the spatial, temporal, and behavioral dimensions of training-related change. Tertiary analyses will address broader motor and cognitive measures as well as participant experience. Exploratory analyses will examine associations between neural and behavioral changes, potential predictors of training response, lesion-related factors, and the persistence of outcomes at the exploratory three-month follow-up where available. In line with the objectives, training-related changes are expected to be reflected in altered resting-state EEG coherence, modulation of MI-related ERD/ERS and hemispheric lateralization, and improvements in upper-limb motor outcomes, with exploratory analyses assessing whether neural changes are associated with behavioral improvements. Due to the longitudinal and multi-modal nature of the study, incomplete datasets may occur, particularly for MRI measurements 44 (e.g., positioning constraints, excessive movement, discomfort, scheduling constraints), camera data acquired at home, or data from the exploratory three-month follow-up assessment. Missing data will not be imputed. Instead, analyses will be performed on available complete cases for each outcome domain. The number of observations contributing to each analysis will be reported transparently. This strategy ensures that all analyses are based on empirically observed data while preserving the integrity of the MBD and minimizing bias introduced by data imputation.

#### 2.6.1 Statistical Methods

Statistical analyses will be adapted to the achieved sample size and the structure of the MBD. Because the primary outcomes of this study (resting-state EEG coherence, MI-related EEG activity) and the MBD-embedded secondary outcome (grip strength) are derived from data collected within the MBD, statistical analysis will first focus on these variables before turning to outcomes assessed at discrete pre- and post-intervention time points.

For these MBD-derived outcomes, analyses will be performed using nonparametric randomization tests in accordance with MBD methodology (74, 75). Randomization tests make use of the fact that the timing of intervention onset was assigned randomly for each participant. For each participant, the observed difference between baseline and intervention phases is compared against the differences that would have resulted if intervention onset had instead occurred at any of the other time points permitted by the randomization scheme, with the actual baseline and intervention data otherwise held identical. This comparison yields a set of possible alternative differences alongside the observed one. The p-value is calculated as the proportion of these alternative differences that are equal to or larger than the observed difference: a small proportion indicates that the observed change is unlikely to have occurred simply by chance at the actual point of intervention onset, supporting the conclusion that the change is intervention-related rather than attributable to general time-related effects (34). As the test statistic, the mean difference between the baseline and intervention phase will be used; given that improvement trends within the baseline phase are plausible in a stroke rehabilitation population, the difference in within-phase slopes between baseline and intervention will additionally be considered as a complementary test statistic, allowing pre-existing trends to be distinguished from intervention-related changes rather than relying solely on phase mean comparisons. These randomization-based analyses include all available datasets and, owing to the triplet structure of the MBD, can additionally be conducted at the level of complete participant triplets (lesion-location group), where the staggered intervention onset across participants is used to distinguish intervention-related changes from general time effects or unspecific session-to-session fluctuations. In addition to randomization-based analyses, individual trajectories will be inspected visually using established single-case design criteria, including level, trend/slope, variability, overlap, immediacy of change, and consistency of effects across tiers (29). Baseline variability and trends are therefore not treated as confounds to be eliminated prior to analysis, but are explicitly incorporated into the statistical and visual interpretation of the MBD outcomes. Unrestricted pairwise comparisons between all individual sessions will not be performed. If the planned sample size is not reached, inferential statistical analyses will be restricted to the level of complete participant triplets and nonparametric randomization tests. All other outcome measures will be reported descriptively in this case.

Secondary outcomes assessed at discrete pre- and post-intervention time points will be evaluated using paired statistical tests (e.g., paired *t*-tests for outcomes assessed at two time points) or repeated-measures analyses of variance (ANOVA) for outcomes assessed at more than two time points, provided the planned sample size (*N* = 21) is reached. For outcomes assessed at both Pre-I and Pre-II, the mean of the two pre-assessment values will be used as the baseline value for pre-post comparisons. Follow-up measurements, where available, will be analyzed exploratorily to assess the persistence of potential effects. For outcomes requiring normalization to enable comparability across participants or sessions, normalization procedures will be applied consistently across all time points within each participant and will be reported for the respective analyses. Depending on the data type, this may include normalization relative to baseline or pre-intervention values, transformation of skewed variables, or modality-specific scaling procedures. Assumptions of parametric tests will be assessed prior to analysis. If assumptions are not met, appropriate non-parametric alternatives will be used, such as Wilcoxon signed-rank tests for paired comparisons or Friedman tests for repeated-measures comparisons. To reduce the risk of inflated type I error due to repeated testing, post-hoc analyses will not be conducted as unrestricted pairwise comparisons between all individual sessions. Where post-hoc comparisons are required, they will be limited to predefined or theoretically justified contrasts and corrected for multiple comparisons where applicable.

Participant experience and related study conduct indicators will be analyzed descriptively. Participant retention, adherence to the planned training schedule, and completion of assessments and training sessions will be reported as proportions. Responses to the study completion questionnaire will be summarized using descriptive statistics.

Correlation analyses will be used to examine associations between changes in neural measures and motor outcomes. Depending on data distribution, Pearson or Spearman correlation coefficients will be reported. Exploratory regression analyses (e.g., stepwise regression) may be conducted to investigate potential predictors of training response, provided that sample size and data structure permit reliable estimation.

All statistical analyses will be conducted using established software and programming environments (e.g., MATLAB, SPSS, R), with significance thresholds reported for each analysis. Given the small sample size and the exploratory character of several analyses, results will be interpreted cautiously, with emphasis on effect sizes, confidence intervals, visual inspection of individual trajectories, and consistency across measures rather than sole reliance on *p*-values.

### 2.7 Data Management

The results of motor and cognitive assessments are recorded on paper during study sessions and subsequently digitized. To ensure data accuracy, digitized data are independently verified by a second person. During MI-NF sessions, behavioral observations and grip-force measurements are recorded analog and later digitized. Paper-based records are compiled per participant and stored in organized folders within a locked cabinet. After completion of data collection, these records are archived in anonymized form and retained for 10 years. MRI data are acquired as DICOM files. EEG recordings are stored in “.xdf”, “.ov”, or “.gdf” formats depending on the recording system. IMU data are saved in “.bin” and “.xml” formats. Camera recordings are saved in “.mp4” format for home-based monitoring and in “.mkv” format for recordings during motor assessments. Digital data are initially stored locally on the recording devices and transferred to a university computer after each session. Subsequently, all data are uploaded to a secure university server. To ensure data integrity and prevent data loss or accidental overwriting, server-stored data are regularly backed up.

#### 2.7.1 Data Monitoring

A formal Data Monitoring Committee was not planned for this study. The intervention is non-invasive and poses minimal risk, and no interim safety or efficacy analyses were required. Data integrity and participant safety are ensured through internal monitoring procedures conducted by the research team.

#### 2.7.2 Data Protection

The data collected in this study are protected under strict confidentiality and data protection regulations. In accordance with the University of Oldenburg’s data protection guidelines, data security and confidentiality are ensured through initial pseudonymization using a unique participant code, followed by full anonymization after completion of data collection. Participants are only identifiable by pseudonyms linked to their personal information through a secure coding list. Access to this list is strictly limited to authorized study personnel. Participants may request deletion of their personal data until anonymization is completed. Personal data, the coding list, and research data are stored separately, with strongly restricted access. Video recordings are stored exclusively under the participant pseudonym without any direct personal identifiers. Due to the sensitive nature of video material, access is strictly regulated. Once anonymized, all data will be securely stored for 10 years in line with good scientific practice.

#### 2.7.3 Harms

The study procedures do not involve direct physical strain. Since participants are stroke survivors with limited motor capacity, only those who feel physically and mentally capable will participate. Participants are advised to stop any activity if overexertion occurs. Repeated mental training may cause concentration-related fatigue, varying individually, and short breaks will be available as needed. Participants can withdraw from the study at any time without explanation.

EEG caps and IMU sensors may cause slight pressure marks on the skin, and conductive gel may, in rare cases, cause mild skin redness that typically fades within minutes or hours. MRI poses minimal risk for participants without metal implants or claustrophobia, though some may find the experience uncomfortable. No risks or side effects are expected from MI-NF training.

#### 2.7.4 Informedness and Consent

Participants receive a written overview outlining the study’s background, objectives, and procedures before their first appointment. This document explains that participation is voluntary, that they may withdraw at any time without providing reasons, and that they will receive compensation for completed hours. It also details the storage of pseudonymized data, the right to request deletion until anonymization, and the option to approve or decline the use of a home camera. The study begins only after both the participant and the investigator have signed the consent form. Following the final data collection, participants receive a full verbal debriefing on the study design and expectations.

## 3 Discussion

This study protocol describes the design of the MINTS study, a longitudinal, multimodal investigation of home-based motor imagery neurofeedback (MI-NF) training in individuals with chronic stroke. While MI-NF has gained increasing attention as a promising adjunct to classical neurorehabilitation, evidence regarding its underlying neural mechanisms, longitudinal effects, and transfer to everyday motor behavior remains limited. MINTS is designed to address these gaps by integrating neurophysiological, neuroimaging, clinical, and real-world movement assessments within a unified framework.

A central feature of the study is the MBD, which is applied to the continuously assessed dependent variables during the baseline and intervention phases. This design allows the investigation of individual change trajectories while explicitly accounting for intra-individual variability and heterogeneity in impairment profiles. Such designs are particularly well suited for neurorehabilitation research, where recovery processes are highly individualized and sample sizes are often small (25). They are also a pragmatic option in cases where interventions are time and resource intensive and thus unsuited for large-scale randomized controlled trials (76).

Repeated assessments across the baseline and intervention phases enable the identification of temporal patterns in neural and behavioral measures in relation to the onset of MI-NF training. An alternative approach would have been a blinded randomized controlled trial including a sham NF condition, in which feedback is provided independently of the targeted neural activity (77). While randomized controlled trials are considered the gold standard for evaluating intervention efficacy (78) and sham-controlled designs are important in NF research (77, 79), the primary aim of this study is to examine within-subject changes in neural activity over the course of MI-NF training, which does not require a control condition. We acknowledge that the absence of a sham group limits the interpretability of clinical outcomes; however, omitting such a condition allows inclusion of a larger sample for the primary analyses. In addition, sham NF raises practical and ethical challenges in this context, including difficulties in maintaining blinding and the need for prolonged non-contingent feedback. Considering these aspects, we deliberately chose an MBD, enabling the investigation of training-related changes through repeated within-subject measurements.

The multimodal assessment strategy of this study combines mobile and stationary EEG, pre- and post-intervention MRI, standardized clinical motor assessments, and extended movement monitoring during activities of daily living. This approach allows training-related changes to be examined across multiple levels, ranging from task-related oscillatory activity and functional connectivity to clinically relevant motor outcomes and everyday movement behavior. By integrating measures with high temporal resolution, high spatial resolution, and ecological validity, the study aims to link neural plasticity to functional relevance in a manner that is rarely achieved in stroke rehabilitation research. Implementing MI-NF training in participants’ home environments reflects realistic conditions for future clinical application. At the same time, standardized procedures, frequent assessments, and centralized data processing are used to maintain methodological rigor. The study design emphasizes within-subject analyses and modality-specific outcome evaluation, allowing meaningful contributions even when not all assessment modalities are available for every participant.

Overall, the MINTS study is designed to generate insights into MI-NF–related neural plasticity and motor behavior in the chronic stage after stroke. The findings are expected to inform the refinement of individualized NF approaches, support the development of scalable home-based rehabilitation strategies, and contribute to a more nuanced understanding of how training-induced neural changes relate to functional improvements in everyday life.

## Data Availability

Data collection for the study described in this protocol is ongoing. Therefore, no complete study dataset is publicly available at this stage. Following completion of data collection, fully anonymized datasets, excluding video recordings, will be made available to external researchers no later than five years after completion of data collection. During the data collection phase, pseudonymized data may be made available to researchers, students, and doctoral candidates at the University of Oldenburg upon reasonable request.

## 4 Abbreviations

ANOVA: Analysis of variance
ARAT: Action Research Arm Test
BaseL: Classifier for left-hand imagery versus baseline activity
BaseR: Classifier for right-hand imagery versus baseline activity
BCI: Brain-computer interface
BDI-II: Beck Depression Inventory-II
BOLD: Blood oxygen level-dependent
CMRR: Center for Magnetic Resonance Research
CSP: Common spatial patterns
DRKS: German Clinical Trials Register (Deutsches Register Klinischer Studien)
DWI: Diffusion-weighted imaging
EEG: Electroencephalography
EMG: Electromyography
EPI: Echo-planar imaging
ERD: Event-related desynchronization
ERS: Event-related synchronization
FGA: Functional Gait Assessment
FLAIR: Fluid-attenuated inversion recovery
FMA-UE: Fugl-Meyer Assessment for Upper Extremity
fMRI: Functional magnetic resonance imaging
GDPR: General Data Protection Regulation
GS: Grip strength
IMU: Inertial measurement unit
ITI: Inter-trial interval
JHFT: Jebsen-Taylor Hand Function Test
KVIQ: Kinesthetic and Visual Imagery Questionnaire
LR: Classifier for left-hand imagery vs. right-hand imagery
M/C: Motor/cognitive function
MA: Motor attempt
MAS: Modified Ashworth Scale
MBD: Multiple baseline design
ME: Motor execution
MFS: Modified Frenchay Scale
MI: Motor imagery
MINTS: Motor Imagery Neurofeedback Training in Stroke (study acronym)
MoCA: Montreal Cognitive Assessment
MRI: Magnetic resonance imaging
NF: Neurofeedback
RS: Resting-state EEG
SF-12: 12-Item Short Form Health Survey
SIS: Stroke Impact Scale
T1: T1-weighted imaging
TAP-M: Test of Attentional Performance – Mobility Version
TE: Echo time
TI: Inversion time
TR: Repetition time

## 5 Declarations

## 5.1 Ethics Approval and Consent to Participate

The MINTS study was approved by the Commission for Research Impact Assessment and Ethics of the University of Oldenburg (approval number: 2023-15 1) and prospectively registered in the German Clinical Trials Register (DRKS; DRKS00036147) prior to participant enrollment. The study is conducted in accordance with local legislation, institutional requirements, and the principles of the Declaration of Helsinki. All participants provide written informed consent prior to participation and receive a compensation of €12 per hour. Data will be published in anonymized, aggregated form, ensuring that no individual participant can be identified.

## 5.2 Consent for Publication

Not applicable

## 5.3 Availability of Data and Materials

The datasets generated during the current study will not be publicly available at the time of publication of this study protocol, as data collection is ongoing. During the data collection phase, pseudonymized data will be available to researchers, students, and doctoral candidates at the University of Oldenburg upon reasonable request. Following completion of the study, fully anonymized datasets (excluding video recordings) will be made available to external researchers no later than five years after completion of data collection.

## 5.6 Authors’ Contributions

J.D.: Conceptualization, Methodology, Investigation, Project administration, Resources, Software, Formal analysis, Visualization, Writing – original draft, Writing – review & editing. L.B.: Methodology, Investigation, Resources, Software, Formal analysis, Visualization, Writing – review & editing. S.R.: Methodology, Investigation, Resources, Writing – review & editing. A.H.: Conceptualization, Funding acquisition, Supervision, Writing – review & editing. C.K.: Conceptualization, Funding acquisition, Methodology, Supervision, Writing – review & editing.

## 5.7 Acknowledgments

This work was supported by the Neuroimaging Unit of the Carl von Ossietzky Universität Oldenburg funded by grants from the German Research Foundation (3T MRI INST 184/152-1 FUGG and MEG INST 184/148-1 FUGG). The CMRR sequence was kindly provided by the University of Minnesota Center for Magnetic Resonance Research. We thank Dr. Tina Schmitt for her valuable support with MRI setup and protocol implementation, as well as Britta Bruns, Gülsen Yanc, and Katharina Grote for operating the MRI system. We also thank Dr. Christiane Gaus for her neuroradiological assessment of the MRI data and for determining lesion-location group assignments. We are grateful to our research assistants Linn Schwarz, Madina Khalillaeva, Ricarda Reinersmann, and Noah Rickermann for their assistance with study preparation and data collection. We acknowledge the use of OpenAI’s ChatGPT (version 4) for assistance with proofreading and language refinement of this manuscript (https://chat.openai.com/).

## 5.8 Publisher’s note

All claims expressed in this article are solely those of the authors and do not necessarily represent those of their affiliated organizations, or those of the publisher, the editors and the reviewers. Any product that may be evaluated in this article, or claim that may be made by its manufacturer, is not guaranteed or endorsed by the publisher.

## Footnotes

1 In exceptional cases where home-based training is not feasible or not desired, sessions may be conducted in a comparable alternative environment (e.g., workplace or a quiet testing room at the university). For simplicity, the term “home-based” is used throughout the manuscript.

